# Baricitinib plus Standard of Care for Hospitalised Adults with COVID-19 on Invasive Mechanical Ventilation or Extracorporeal Membrane Oxygenation: Results of a Randomised, Placebo-Controlled Trial

**DOI:** 10.1101/2021.10.11.21263897

**Authors:** E. Wesley Ely, Athimalaipet V. Ramanan, Cynthia E. Kartman, Stephanie de Bono, Ran Liao, Maria Lucia B. Piruzeli, Jason D. Goldman, José Francisco Kerr Saraiva, Sujatro Chakladar, Vincent C. Marconi, on behalf of the COV-BARRIER Study Group

**Affiliations:** Critical Illness, Brain Dysfunction, and Survivorship (CIBS) Center, Division of Allergy, Pulmonary, and Critical Care Medicine, Department of Medicine at Vanderbilt University Medical Center, Nashville, TN, USA; Tennessee Valley Veteran’s Affairs Geriatric Research Education Clinical Center (GRECC), Nashville, TN, USA; Translational Health Sciences, University of Bristol, Bristol, UK; Department of Paediatric Rheumatology, Bristol Royal Hospital for Children, Bristol, UK; Eli Lilly and Company, Indianapolis, IN, USA; Swedish Center for Research and Innovation, Swedish Medical Center, Providence St. Joseph Health Seattle, WA, USA; Division of Allergy and Infectious Disease, Department of Medicine, University of Washington, Seattle, WA, USA; Instituto de Pesquisa Clínica de Campinas (IPECC), Campinas, SP, Brazil; Emory University School of Medicine, Rollins School of Public Health, Emory Vaccine Center Atlanta, GA, USA; Atlanta Veterans Affairs Medical Center, Decatur, GA, USA

## Abstract

**Background:** The oral, selective Janus kinase (JAK)1/JAK2 inhibitor baricitinib demonstrated efficacy in hospitalised adults with COVID-19. This study evaluates the efficacy and safety of baricitinib in critically ill adults with COVID-19 requiring invasive mechanical ventilation (IMV) or extracorporeal membrane oxygenation (ECMO).

**Methods:** COV-BARRIER was a global, phase 3, randomised, double-blind, placebo-controlled trial in patients with confirmed SARS-CoV-2 infection (ClinicalTrials.gov NCT04421027). This addendum trial added a critically ill cohort not included in the main COV-BARRIER trial. Participants on baseline IMV/ECMO were randomly assigned 1:1 to baricitinib 4-mg (n=51) or placebo (n=50) for up to 14 days in combination with standard of care (SOC). Prespecified endpoints included all-cause mortality through days 28 and 60, and number of ventilator-free days, duration of hospitalisation, and time to recovery through day 28. Efficacy and safety analyses included the intent-to-treat and safety populations, respectively.

**Findings:** SOC included baseline systemic corticosteroid use in 86% of participants. Treatment with baricitinib significantly reduced 28-day all-cause mortality compared to placebo (39·2% vs 58·0%; hazard ratio [HR]=0·54 [95%CI 0·31–0·96]; p=0·030). One additional death was prevented for every six baricitinib-treated participants. Significant reduction in 60-day mortality was also observed (45·1% vs 62·0%; HR=0·56 [95%CI 0·33–0·97]; p=0·027).

Baricitinib-treated participants showed numerically more ventilator-free days (8.1 vs 5.5 days, p=0.21) and spent over 2 days less in the hospital than placebo-treated participants (23·7 vs 26·1 days, p=0·050). The rates of infections, blood clots, and adverse cardiovascular events were similar between treatment arms.

**Interpretation:** In critically ill patients with COVID-19 already receiving IMV/ECMO, treatment with baricitinib as compared to placebo (in combination with SOC, including corticosteroids) showed mortality HR of 0·56, corresponding to a 44% relative reduction at 60 days. This is consistent with the mortality reduction observed in less severely ill hospitalised primary COV-BARRIER study population.

**Funding:** Eli Lilly and Company.

**Research in context:** *Evidence before this study:* We evaluated current and prior studies assessing the efficacy and safety of interventions in patients requiring invasive mechanical ventilation (IMV) and searched current PubMed using the terms “COVID-19”, “SARS-CoV-2”, “treatment”, “critical illness”, “invasive mechanical ventilation”, “baricitinib”, and “JAK inhibitor” for articles in English, published until December 1, 2020, regardless of article type. We also reviewed the NIH and IDSA COVID-19 guidelines and reviewed similar terms on clinicaltrials.gov. When the critical illness addendum study to COV-BARRIER study was designed, there was only one open-label study of dexamethasone showing mortality benefit in hospitalised patients with COVID-19 requiring IMV. Small studies of interleukin-6 inhibitors had shown no effect and larger trials were underway. Guidelines recommended use of dexamethasone with or without remdesivir and recommended against the use of interleukin-6 inhibitors, except in a clinical trial. Overall, there were no reported double-blind, placebo-controlled phase 3 trials which included corticosteroids as part of SOC investigating the efficacy and safety of novel treatments in the NIAID-OS 7 population. Baricitinib’s mechanism of action as a JAK1 and JAK2 inhibitor was identified as a potential intervention for the treatment of COVID-19 given its known anti-cytokine properties and potential antiviral mechanism for targeting host proteins mediating viral endocytosis Data from the NIAID sponsored ACTT-2 trial showed that baricitinib when added to remdesivir improved time to recovery and other outcomes including mortality compared to placebo plus remdesivir. A numerically larger proportion of participants who received baricitinib plus remdesivir showed an improvement in ordinal scale compared to those who received placebo plus remdesivir at day 15 in participants requiring IMV (NIAID-OS score of 7) at baseline. We designed COV-BARRIER, a phase 3, global, double-blind, randomised, placebo-controlled trial, to evaluate the efficacy and safety of baricitinib in combination with SOC (including corticosteroids) for the treatment of hospitalised adults with COVID-19 who did not require mechanical ventilation (i.e., NIAID-OS 4-6). A significant reduction in mortality was found after 28 days between baricitinib and placebo (HR 0·57, corresponding to a 43% relative reduction, p=0·0018); one additional death was prevented per 20 baricitinib-treated participants. In the more severely ill NIAID-OS 6 subgroup, one additional death was prevented per nine baricitinib-treated participants (HR 0·52, corresponding to a 48% relative reduction, p=0·0065). We therefore implemented an addendum to the COV-BARRIER trial to evaluate the benefit/risk of baricitinib in the critically ill NIAID-OS 7 population and considered the sample size of 100 participants sufficient for this trial.

*Added value of this study:* This was the first phase 3 study to evaluate baricitinib in addition to the current standard of care (SOC), including antivirals, anticoagulants, and corticosteroids, in patients who were receiving IMV or extracorporeal membrane oxygenation at enrolment. This was a multinational, randomised, double-blind, placebo-controlled trial in regions with high COVID-19 hospitalisation rates. Treatment with baricitinib reduced 28-day all-cause mortality compared to placebo (HR 0·54, 95% CI 0·31–0·96; nominal p=0·030), corresponding to a 46% relative reduction, and significantly reduced 60-day all-cause mortality (HR 0·56, 95% CI 0·33–0·97; p=0·027); overall, one additional death was prevented per six baricitinib-treated participants. Numerical improvements in endpoints such as number of ventilator-free days, duration of hospitalisation, and time to recovery were demonstrated. The frequency of serious adverse events, serious infections, and venous thromboembolic events was similar between baricitinib and placebo, respectively. The COV-BARRIER study overall trial results plus these COV-BARRIER addendum study data in mechanically ventilated and ECMO patients provide important information in context of other large, phase 3 randomised trials in participants with invasive mechanical ventilation at baseline. The RECOVERY study reported mortality of 29·3% following treatment with dexamethasone compared to 41·4% for usual care (rate ratio of 0·64, corresponding to a 36% relative reduction) and 49% mortality in participants who received tocilizumab compared to 51% for usual care (rate ratio of 0.93, corresponding to a 7% relative reduction). The ACTT-2 study reported 28-day mortality of 23·1% and 22·6% in the baricitinib plus remdesivir and placebo plus remdesivir groups, respectively, in this critically ill patient population; however, the primary outcome of this trial was time to recovery, so was not powered to detect a change in mortality.

*Implications of all the available evidence:* In this phase 3 addendum trial, baricitinib given in addition to SOC (which predominantly included corticosteroids) had a significant effect on mortality reduction by 28 days in critically ill patients, an effect which was maintained by 60 days. These data were comparable with those seen in the COV-BARRIER primary study population of hospitalised patients, but which excluded patients who required IMV or extracorporeal membrane oxygenation at enrolment. These findings suggest that baricitinib has synergistic effects to other SOC treatment modalities including remdesivir and dexamethasone. Based on the available evidence, baricitinib is a novel treatment option to decrease mortality in hospitalised, critically ill patients with COVID-19 even when started late in the disease process after steroids, mechanical ventilation, and ECMO have already been implemented.

## INTRODUCTION

Patients who require hospitalisation following infection with severe acute respiratory syndrome coronavirus 2 (SARS-CoV-2) will often experience an intense hyperinflammatory state that may lead to multiple organ dysfunction including acute respiratory distress syndrome, septic shock, and death.^1-4^ There have been many recent treatment advances in therapeutics for patients hospitalised with coronavirus disease 19 (COVID-19) such as remdesivir, dexamethasone, tocilizumab, and baricitinib.^6-11^ Globally, however, there remains a critical and urgent need to evaluate new treatment options to reduce mortality in hospitalised patients with COVID-19 due to the high occurrence of deaths which persists despite these improvements in standard of care (SOC). For example, the World Health Organisation reported that over 22,000 deaths were confirmed in the region of the Americas in the week from August 29 to September 04, 2021. Mortality remains particularly high amongst critically ill patients who require invasive mechanical ventilation (IMV) or extracorporeal membrane oxygenation (ECMO), which is the primary means of treatment in patients with severe COVID-19, with an estimated case fatality rate of 45% in this population.^5^ In the platform RECOVERY trial, mortality was 29·3% in persons with baseline IMV randomised to dexamethasone compared to 41·4% for usual care, with the rate ratio of 0·64 (95%CI 0·51-0·81) corresponding to a 36% relative reduction in mortality in the IMV subgroup.^8^ Similarly, mortality was 49% in participants with baseline IMV randomised to tocilizumab compared to 51% for usual care with rate ratio 0·93 (95% 0·74−1·18) corresponding to a 7% relative reduction in mortality in that subgroup.^9^ Notably in the tocilizumab study, benefits of mortality across the whole study population were only seen in those with concomitant use of corticosteroids. Thus, interventions to reduce mortality in critically ill patients with COVID-19 remain a crucial unmet medical need.

In February 2020, baricitinib, a selective Janus kinase (JAK)1/JAK2 inhibitor,^12,13^ was identified by an artificial intelligence platform as a potential intervention for the treatment of COVID-19 given its known anti-inflammatory profile in patients with autoimmune diseases^14^ and potential for targeting host proteins for its antiviral mechanism.^15,16^ Several observational studies carried out on small cohorts of hospitalised patients with COVID-19 (including elderly patients) provided the first evidence of clinical improvement associated with baricitinib treatment.^17-20^

The Adaptive COVID-19 Treatment Trial 2 (ACTT-2), a NIH-sponsored global double-blind, randomised, placebo-controlled, phase 3 trial in hospitalised adults with COVID-19, found that baricitinib plus remdesivir was superior to remdesivir in reducing time to recovery (rate ratio for recovery 1·16, 95% CI 1·01–1·32; p=0·03), with a numerical reduction in 28-day mortality reported (5·1% vs 7·8%, hazard ratio [HR] 0·65, 95% CI 0·39–1·09 corresponding to a 35% relative reduction) and fewer serious adverse events (SAEs) in the participants receiving baricitinib plus remdesivir.^7^ In 111 participants enrolled with baseline use of IMV or ECMO, there were no significant differences in 28-day mortality or recovery. However, there was a numerical improvement seen in the key secondary outcome of ordinal score on day 15 for participants who received baricitinib and remdesivir compared to those who received remdesivir plus placebo (odds ratio 1·7, 95% CI 0·8–3·4).

COV-BARRIER was a phase 3, global, double-blind, randomised, placebo-controlled trial designed to evaluate the efficacy and safety of baricitinib in combination with SOC, which could include corticosteroids, for the treatment of hospitalised adults with COVID-19 who did not require mechanical ventilation (i.e., National Institute of Allergy and Infectious Disease Ordinal Scale [NIAID-OS] 4-6). While the primary endpoint of reduction of disease progression did not reach statistical significance, a significant reduction in 28-day all-cause mortality was found between baricitinib and placebo (HR 0·57, 95% CI 0·41–0·78; p=0·0018) corresponding to a 43% relative reduction; one additional death was prevented per 20 baricitinib-treated participants.^11^ The mortality benefits were more pronounced in the more severely ill subgroup who required high flow oxygen or non-invasive ventilation (NIAID-OS 6), with a HR of 0·52 (95% CI 0·33-0·80; nominal p=0·0065) for 28-day all-cause mortality between baricitinib and placebo corresponding to a 48% relative reduction in mortality. In participants with baseline NIAID-OS 6, one additional death was prevented per nine baricitinib-treated participants.

The Food and Drug Administration (FDA) issued an Emergency Use Authorization (EUA) for the use of baricitinib to treat COVID-19 in hospitalised adults and paediatric patients 2 years of age or older requiring supplemental oxygen, non-invasive or invasive mechanical ventilation, or ECMO.^21^ The EUA was first issued in November 2020 with ACTT-2 results and later updated in July 2021 with COV-BARRIER NIAID-OS 4-6 results. Here we report a trial conducted as an addendum to COV-BARRIER trial to investigate the efficacy and safety of baricitinib in combination with SOC for the treatment of adult participants with COVID-19 requiring IMV or ECMO.

## METHODS

### Study Design and participants

The COV-BARRIER addendum trial in persons with baseline IMV/ECMO was a multicentre, randomised, double-blind, placebo-controlled, parallel-group, phase 3 trial which were enrolled from 18 centres in four countries (Argentina, Brazil, Mexico, United States). Participants were enrolled from December 23, 2020 to April 10, 2021. A detailed description of the parent study design has been previously published.^11^

Eligible participants were ≥18 years of age, hospitalised with laboratory-confirmed SARS-CoV-2 infection, use of IMV or ECMO at study entry and randomisation, had evidence of pneumonia or clinical symptoms of COVID-19, and had at least one elevated inflammatory marker above the upper limit of normal range based on the local laboratory result (C-reactive protein, D-dimer, lactate dehydrogenase, or ferritin). Dexamethasone use was permitted as described in the RECOVERY trial,^8^ but patients were excluded if receiving high dose corticosteroids (>20 mg per day [or prednisone equivalent] administered for >14 consecutive days in the month before study entry, unless indicated per SOC for a concurrent condition, such as asthma, chronic obstructive pulmonary disease, or adrenal insufficiency), immunosuppressants, biologics, T cell or B cell-targeted therapies, interferon, or JAK inhibitors; receipt of convalescent plasma or intravenous immunoglobulin for COVID-19; or suspected serious active bacterial, fungal, or other infection, or untreated tuberculosis infection. All participants received SOC in keeping with local clinical practice for COVID-19 management, which could include corticosteroids, antivirals, and/or other treatments. Prophylaxis for venous thromboembolic events (VTE) per local practice was required for all participants unless contraindicated.

COV-BARRIER was conducted in accordance with ethical principles of the Declaration of Helsinki and Good Clinical Practice guidelines. All sites received approval from the authorized institutional review board. All participants (or legally authorized representatives) provided informed consent.

### Procedures

Participants who met all criteria for enrolment were randomised 1:1 to baricitinib or matched placebo at day 1. Baricitinib 4-mg or placebo was crushed for nasogastric tube delivery (or given orally when feasible) and given daily for up to 14 days or until discharge from hospital, whichever occurred first. Participants randomised to baricitinib with baseline eGFR ≥30 to <60 mL/min/1·73 m^2^ received baricitinib 2-mg or placebo to match. If eGFR decreased to ≥30 to <60 mL/min/1·73 m^2^ after randomisation, patients received baricitinib 2-mg until eGFR returned to ≥60 mL/min/1·73 m^2^.

For efficacy and health outcomes, baseline measurements were defined as the last non-missing assessment recorded on, or prior to, the first study drug administration at study day 1. Efficacy and safety were evaluated for all participants up to day 28; all-cause mortality was also evaluated up to day 60. Participants had a follow-up visit ∼28 days after receiving their last dose of study drug. Participant disposition with reasons for discontinuation which include adverse events (AEs) (including death) are detailed in Figure 1.

**Figure 1.**
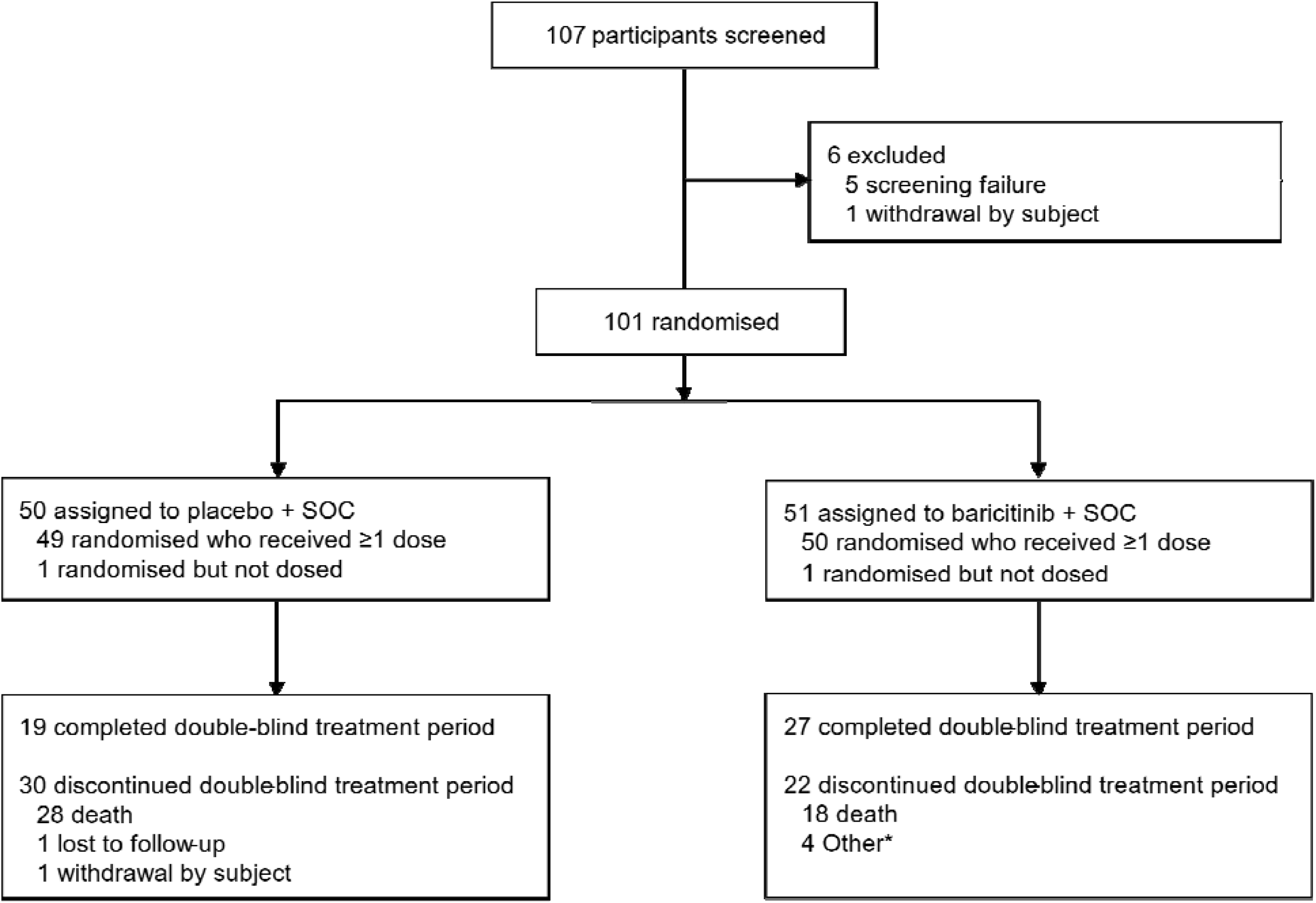
Trial profile. *4 participants with discontinuation status recorded as ‘other’ discontinued due to transfer to another hospital.

### Outcomes

The prespecified endpoints included all-cause mortality by day 28 and day 60; number of ventilator-free days; overall improvement (assessed by odds of improvement in clinical status) on National Institute of Allergy and Infectious Disease Ordinal Scale (NIAID-OS) evaluated at days 4, 7, 10, 14, and 28; proportion of participants with at least 1-point improvement on the NIAID-OS or live discharge from the hospital at days 4, 7, 10, 14, and 28; duration of hospitalisation; and time to recovery through day 28. As the cohort reported here was an addendum, all endpoints are considered exploratory.

### Statistical analyses

In this analysis, the sample size for this cohort was approximately 50 participants per treatment arm, and was selected to match the number of participants enrolled in the ACTT-2 study requiring IMV or ECMO at baseline. This was deemed a sufficient sample size to evaluate participants treated with baricitinib in addition to SOC in a randomised controlled study who were hospitalised and on IMV or ECMO at baseline.

Efficacy data were analysed with the intent-to-treat population, defined as all randomised participants. Patients who discontinued on the day of randomisation with no baseline or post-baseline OS data were excluded. Log-rank test and HR from Cox proportional hazard model were used for time-to-event analyses. Logistic regression with the last observation carried forward methodology was used for dichotomous endpoints, proportional odds model was used for ordinal endpoints, and analysis of variance model was used for continuous endpoints. These statistical models were adjusted for treatment, baseline age group (<65, ≥65 years), and region. Analysis of NIAID-OS outcomes at day 60 was carried out using descriptive statistics on observed values. Safety analyses included all randomised participants who received ≥1 dose of study drug and who did not discontinue the study for the reason of ‘lost to follow-up’ at the first post-baseline visit. AEs were inclusive of the 28-day treatment period. Statistical tests of treatment effects were performed at a 2-sided significance level of 0·05, unless otherwise stated. Statistical analyses were performed using SAS® Version 9·4 or higher or R. This trial is registered with ClinicalTrials.gov, NCT04421027.

## RESULTS

Between December 23, 2020 and April 10, 2021, 101 participants were randomly assigned to receive once-daily baricitinib 4-mg plus SOC (n=51) or placebo plus SOC (n=50); 46 (46·5%) participants completed the 28-day treatment period (Figure 1). Of the 52 participants (52·5%) who discontinued from the treatment period, 46 (88·5%) discontinuations were due to death. No randomly assigned participants were excluded from the intent-to-treat population.

Baseline demographics and disease characteristics were balanced among treatment groups. The mean age of the participants was 58·6 years (SD 13·8), 55 (54·5%) participants were male (Table 1). The trial was carried out in four countries: Mexico (31 participants, 30·7%), Brazil (29 participants, 28·7%), Argentina (21 participants, 20·8%), and United States (20 participants, 19·8%). Overall, 32 participants (32·3%) were American Indian or Alaska Native, one (1·0%) was Asian, two (2·0%) were Black or African American, and 62 (62·6%) were white. The majority (93 [93·9%]) of participants had symptoms ≥7 days prior to enrolment. At baseline, 87 (86·1%) participants received systemic corticosteroids; two (2·0%) participants received remdesivir (Table 1). All participants (100%) had ≥1 pre-existing comorbidity of interest.

**Table 1:** Baseline demographics and disease characteristics.

|  | **Baricitinib 4-mg** |  |  |
| --- | --- | --- | --- |
|  | **Placebo + SOC** | **+ SOC** | **All Participants** |
|  | **(N= 50)** | **(N= 51)** | **(N=101)** |
| Age, years | 58.8 (15.2) | 58.4 (12.4) | 58.6 (13.8) |
| Sex |  |  |  |
| Male | 30 (60.0) | 25 (49.0) | 55 (54.5) |
| Female | 20 (40.0) | 26 (51.0) | 46 (45.5) |
| Race |  |  |  |
| American Indian or Alaska Native* | 17 (34.7) | 15 (30.0) | 32 (32.3) |
| Asian | 1 (2.0) | 0 (0.0) | 1 (1.0) |
| Black or African American | 1 (2.0) | 1 (2.0) | 2 (2.0) |
| Multiple | 0 | 2 (4.0) | 2 (2.0) |
| White | 30 (61.2) | 32 (64.0) | 62 (62.6) |
| Missing | 1 | 1 | 2 |
| Country |  |  |  |
| Argentina | 9 (18.0) | 12 (23.5) | 21 (20.8) |
| Brazil | 14 (28.0) | 15 (29.4) | 29 (28.7) |
| Mexico | 17 (34.0) | 14 (27.5) | 31 (30.7) |
| United States (incl Puerto Rico) | 10 (20.0) | 10 (19.6) | 20 (19.8) |
| BMI | 32.1 (6.3) | 34.3 (7.8) | 33.2 (7.1) |
| Disease duration of symptoms prior to enrolment |  |  |  |
| <7 days | 4 (8.3) | 2 (3.9) | 6 (6.1) |
| ≥7 days | 44 (91.7) | 49 (96.1) | 93 (93.9) |

|  | Baricitinib 4-mg |  |  |
| --- | --- | --- | --- |
|  | Placebo + SOC<br>(N= 50) | + SOC<br>(N= 51) | All Participants<br>(N=101) |
| Key Concomitant Medications at baseline |  |  |  |
| Remdesivir use | 2 (4.0) | 0 (0.0) | 2 (2.0) |
| Corticosteroid use | 44 (88.0) | 43 (84.3) | 87 (86.1) |
| Pre-existing Comorbid Conditions of Interest† |  |  |  |
| Obesity | 29 (58.0) | 28 (54.9) | 57 (56.4) |
| Diabetes (Type I and Type II) | 16 (32.0) | 20 (39.2) | 36 (35.6) |
| Chronic respiratory disease | 2 (4.0) | 1 (2.0) | 3 (3.0) |
| Hypertension | 24 (48.0) | 31 (60.8) | 55 (54.5) |
| ECMO at baseline | 1 (2.0) | 3 (5.9) | 4 (4.0) |
| Inflammatory Markers, median |  |  |  |
| CRP (mg/L) | 109.5 | 124.9 | 115.4 |
| D-dimer (mg/L) | 1.6 | 1.6 | 1.6 |
| Lactate dehydrogenase (U/L) | 543.6 | 499.5 | 531.5 |
| Ferritin (pmol/L) | 2836.9 | 2622.0 | 2711.0 |

Data are mean (SD) or n (%). CRP=C-reactive protein. ECMO= extracorporeal membrane oxygenation. N=number of patients in the analysis population. n=number of participants in the specified category. SD=standard deviation. SOC=standard of care. \*Includes participants of Hispanic descent. †Patients with estimated glomerular filtration rate <30 mL/min/1.73 m<sup>2</sup> were excluded from study enrolment.

Baseline C-reactive protein was elevated with a median value of 115·4 mg/L in the overall study population and was balanced between baricitinib and placebo groups (124·9 mg/L and 109·5 mg/L, respectively). VTE prophylaxis was utilised as required, the majority of participants utilised enoxaparin (78·2%). The median number of days of treatment exposure was 12·0 (interquartile rage [IQR] 5.0–14.0) in the placebo group and 11·0 (IQR 7·0–14·0) in the baricitinib group.

Treatment with baricitinib significantly reduced deaths, as measured by all-cause mortality. At day 28, mortality was 39·2% (20 of 51) in the baricitinib group and 58·0% (29 of 50) in the placebo group (HR 0·54 [95% CI 0·31–0·96]; nominal p=0·030), corresponding to a 46% relative reduction; there was an absolute risk reduction of 18·8 percentage points and overall, one additional death was prevented per six baricitinib-treated participants (Table 2, Figure 2A). Between day 28 and day 60, five additional deaths occurred in the overall population with a similar number of events in the baricitinib (n=3) and placebo (n=2) groups. The 60-day mortality remained significantly lower in the baricitinib group compared to the placebo group (45·1% [23/51] vs 62·0% [31/50]; HR 0·56 [95% CI 0·33–0·97]; p=0·027), corresponding to a 44% relative reduction and an absolute risk reduction of 16·9 percentage points (Figure 2B).

**Table 2:**
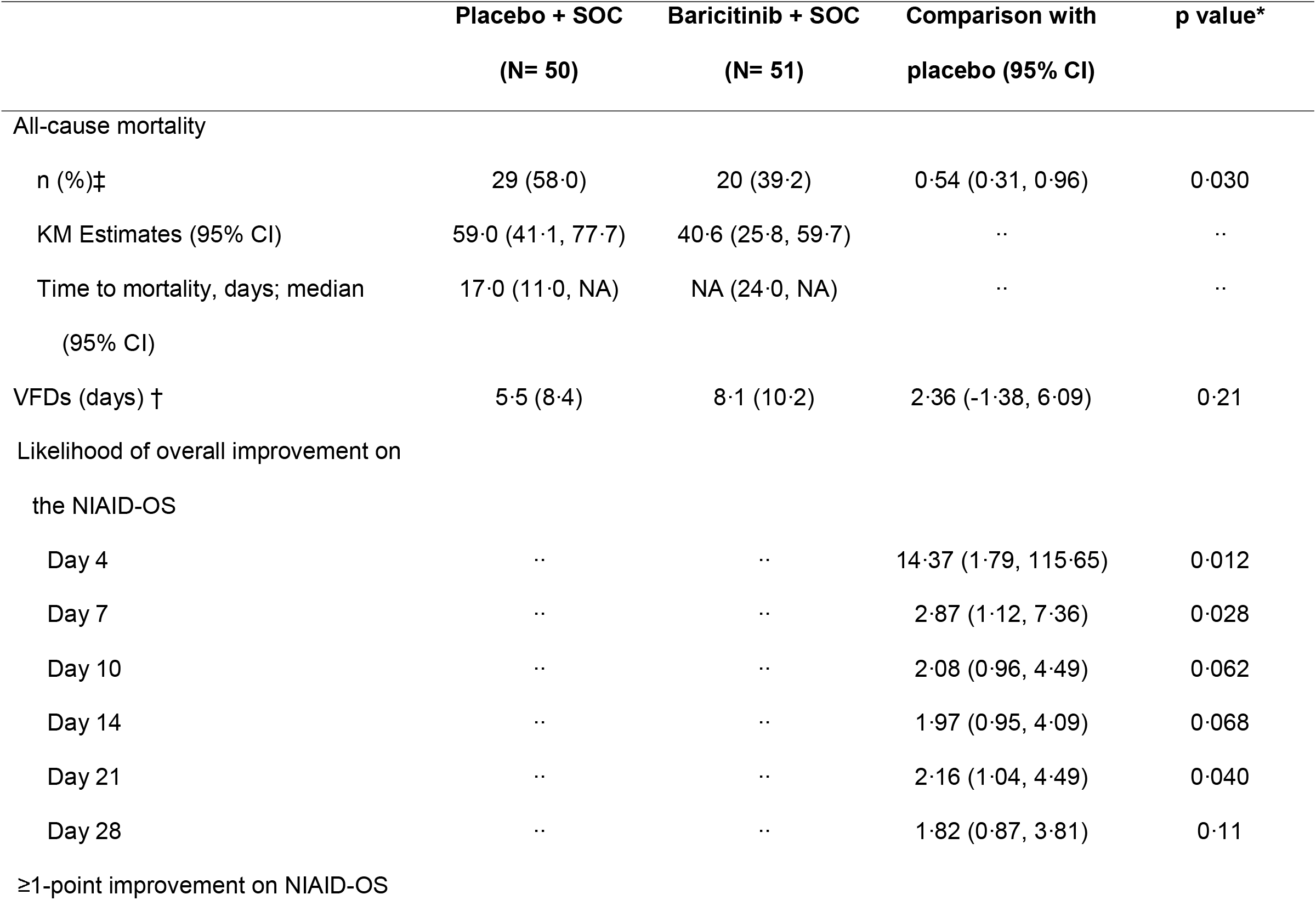
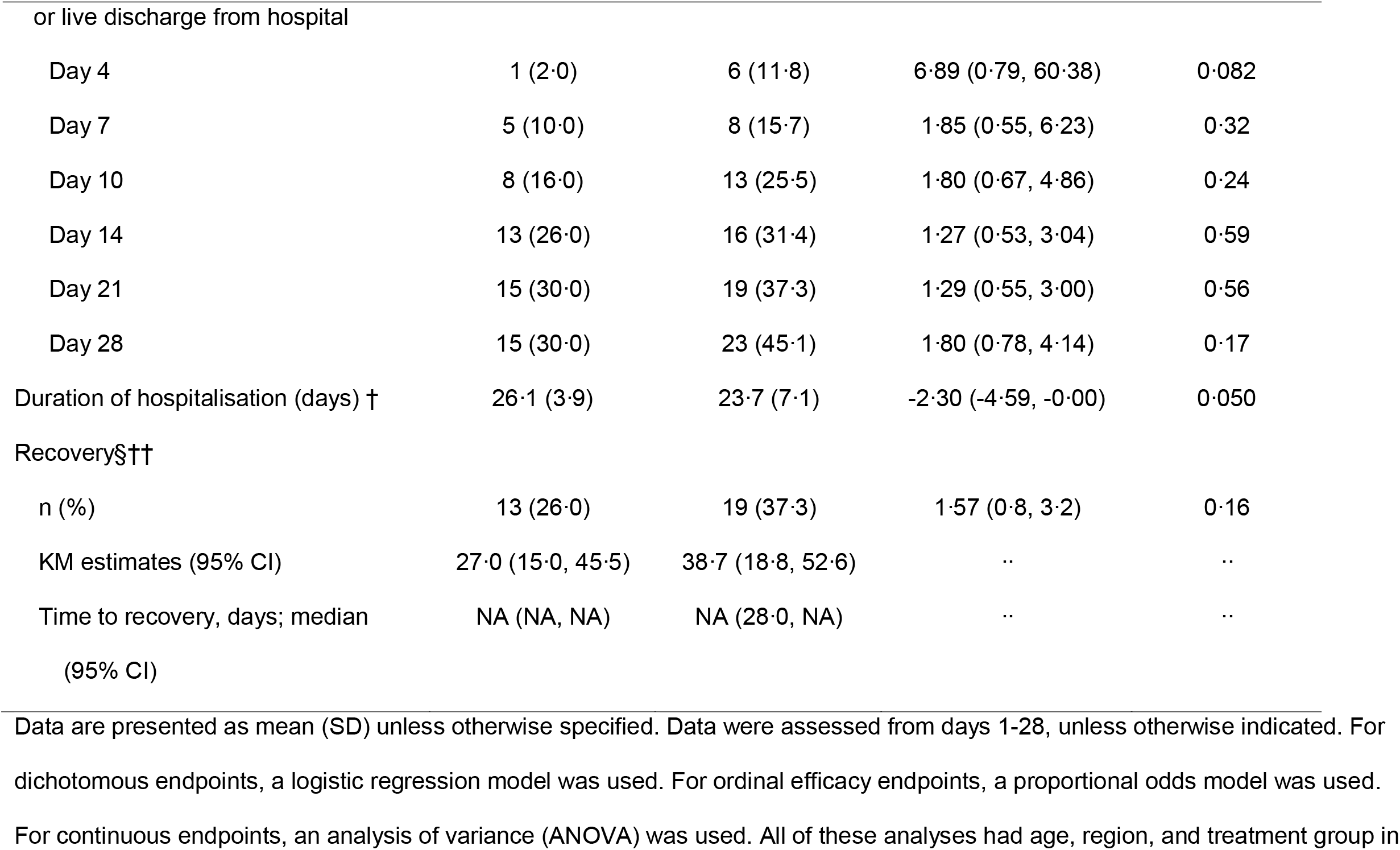
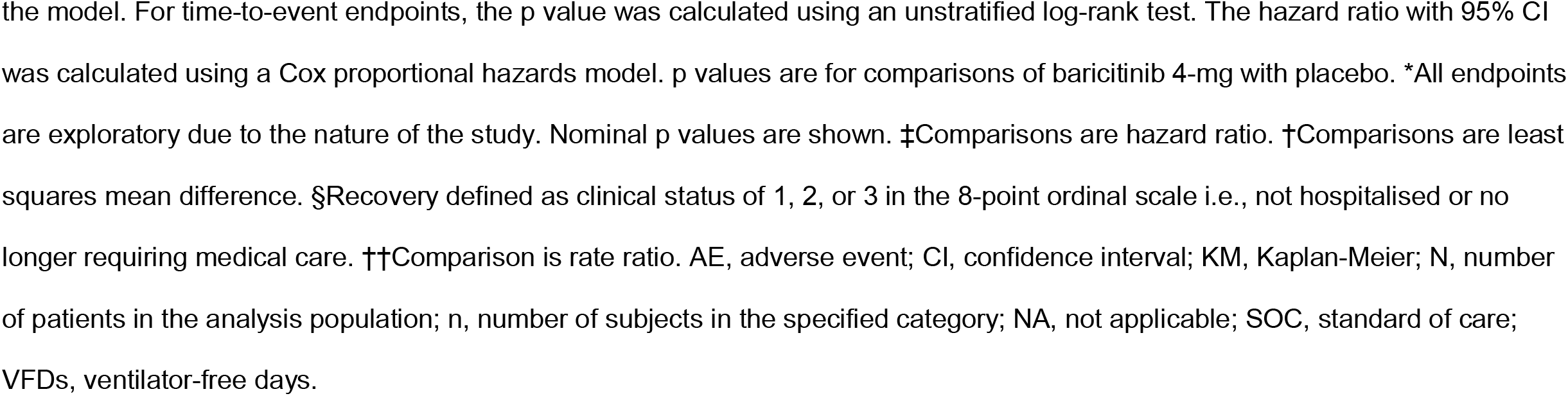
Overview of efficacy outcomes in the intent-to-treat population by day 28.

|  | Placebo + SOC<br>(N= 50) | Baricitinib + SOC<br>(N= 51) | Comparison with<br>placebo (95% CI) | p value* |
| --- | --- | --- | --- | --- |
| All-cause mortality |  |  |  |  |
| n (%)‡ | 29 (58.0) | 20 (39.2) | 0.54 (0.31, 0.96) | 0.030 |
| KM Estimates (95% CI) | 59.0 (41.1, 77.7) | 40.6 (25.8, 59.7) | .. | .. |
| Time to mortality, days; median<br>(95% CI) | 17.0 (11.0, NA) | NA (24.0, NA) | .. | .. |
| VFDs (days) † | 5.5 (8.4) | 8.1 (10.2) | 2.36 (-1.38, 6.09) | 0.21 |
| Likelihood of overall improvement on<br>the NIAID-OS |  |  |  |  |
| Day 4 | .. | .. | 14.37 (1.79, 115.65) | 0.012 |
| Day 7 | .. | .. | 2.87 (1.12, 7.36) | 0.028 |
| Day 10 | .. | .. | 2.08 (0.96, 4.49) | 0.062 |
| Day 14 | .. | .. | 1.97 (0.95, 4.09) | 0.068 |
| Day 21 | .. | .. | 2.16 (1.04, 4.49) | 0.040 |
| Day 28 | .. | .. | 1.82 (0.87, 3.81) | 0.11 |
| ≥1-point improvement on NIAID-OS |  |  |  |  |
| or live discharge from hospital |  |  |  |  |
| Day 4 | 1 (2.0) | 6 (11.8) | 6.89 (0.79, 60.38) | 0.082 |
| Day 7 | 5 (10.0) | 8 (15.7) | 1.85 (0.55, 6.23) | 0.32 |
| Day 10 | 8 (16.0) | 13 (25.5) | 1.80 (0.67, 4.86) | 0.24 |
| Day 14 | 13 (26.0) | 16 (31.4) | 1.27 (0.53, 3.04) | 0.59 |
| Day 21 | 15 (30.0) | 19 (37.3) | 1.29 (0.55, 3.00) | 0.56 |
| Day 28 | 15 (30.0) | 23 (45.1) | 1.80 (0.78, 4.14) | 0.17 |
| Duration of hospitalisation (days) † | 26.1 (3.9) | 23.7 (7.1) | -2.30 (-4.59, -0.00) | 0.050 |
| Recovery§†† |  |  |  |  |
| n (%) | 13 (26.0) | 19 (37.3) | 1.57 (0.8, 3.2) | 0.16 |
| KM estimates (95% CI) | 27.0 (15.0, 45.5) | 38.7 (18.8, 52.6) | .. | .. |
| Time to recovery, days; median<br>(95% CI) | NA (NA, NA) | NA (28.0, NA) | .. | .. |

Data are presented as mean (SD) unless otherwise specified. Data were assessed from days 1-28, unless otherwise indicated. For dichotomous endpoints, a logistic regression model was used. For ordinal efficacy endpoints, a proportional odds model was used. For continuous endpoints, an analysis of variance (ANOVA) was used. All of these analyses had age, region, and treatment group in
the model. For time-to-event endpoints, the p value was calculated using an unstratified log-rank test. The hazard ratio with 95% CI was calculated using a Cox proportional hazards model. p values are for comparisons of baricitinib 4-mg with placebo. \*All endpoints are exploratory due to the nature of the study. Nominal p values are shown. ‡Comparisons are hazard ratio. †Comparisons are least squares mean difference. §Recovery defined as clinical status of 1, 2, or 3 in the 8-point ordinal scale i.e., not hospitalised or no longer requiring medical care. ††Comparison is rate ratio. AE, adverse event; CI, confidence interval; KM, Kaplan-Meier; N, number of patients in the analysis population; n, number of subjects in the specified category; NA, not applicable; SOC, standard of care; VFDs, ventilator-free days.

**Figure 2:**
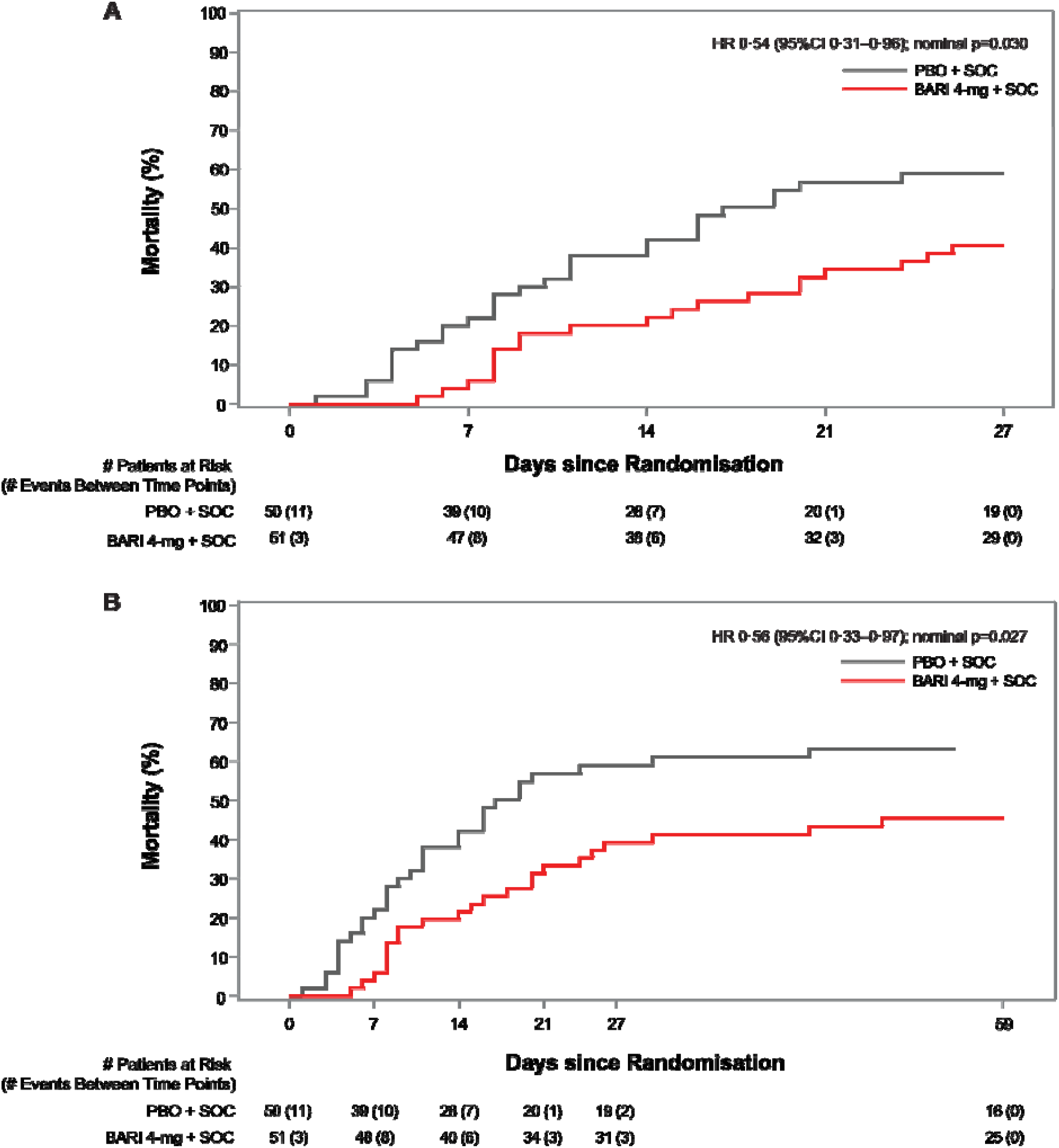
Kaplan-Meier estimates of all-cause mortality (including potentially related with COVID-19 and attributed to adverse events) at Day 28 (A) and Day 60 (B). The numbers at risk at Days 27 and 59 represent the numbers of participants with available data at Days 28 and 60, respectively. *a HR and 95% CIs were calculated using cox proportional hazard regression model adjusted for age (<65 years, >=65 years), region (United States, rest of world); unstratified. *b p value was calculated from unstratified log-rank test. BARI, baricitinib; CI, confidence interval; HR, hazard ratio; NA, not applicable; PBO, placebo; SOC, standard of care.

Patients treated with baricitinib also showed numerical improvements compared to those treated with placebo in other exploratory endpoints examined in this addendum trial. These endpoints included a numerical increase in the number of ventilator-free days (mean 8·1 days vs 5·5 days, p=0·21), overall improvement in NIAID-OS (likelihood of improvement at day 28 odds ratio 1·82 [0·87, 3·81], p=0·11), proportion having ≥1-point improvement on NIAID-OS or hospital discharge (45·1% [n=23] for baricitinib vs 30·0% [n=15] for placebo at day 28, p=0·17), and shorter duration of hospitalisation (23·7 days vs 26·1 days, p=0·050). More patients recovered in the baricitinib group compared to placebo by day 28 (19 [37·3%] vs 13 [26·0%]). Additionally, a numerically higher proportion of participants recovered (reached NIAID-OS 1 or 2) at day 60 in the baricitinib group compared to placebo group, 47·1% vs 32·0%, respectively (Table S7). Outcomes are shown in more detail in Table 2, and analysis of outcomes by subgroup (baseline corticosteroid use, baseline remdesivir use, and country) at day 28 and day 60 are given in the supplemental appendix (Tables S1–S7).

The proportion of participants with ≥1 treatment-emergent AE (TEAE) was 88·0% [44 of 50] in the baricitinib group and 95·9% [47 of 49] in the placebo group; for SAEs, these proportions were 50·0% (25 of 50) and 71·4% (35 of 49), respectively (Table 3). The number of participants who discontinued study treatment due to AE (14 [28·0%] vs 17 [34·7%]) and the frequency of deaths reported as due to AE (five [10·0%] vs three [6·1%]) was similar in both groups. The number of participants with treatment-emergent infections were comparable across treatment arms (35 [70·0%] in baricitinib and 35 [71·4%] in placebo). Serious infections were reported for 22 (44·0%) participants who received baricitinib and 26 (53·1%) who received placebo. There was a similar distribution of positively adjudicated VTEs (three [6·0%] vs three [6·1%]) with baricitinib and placebo groups, respectively. Safety data are described in more detail in Tables 3 and S8.

**Table 3.**
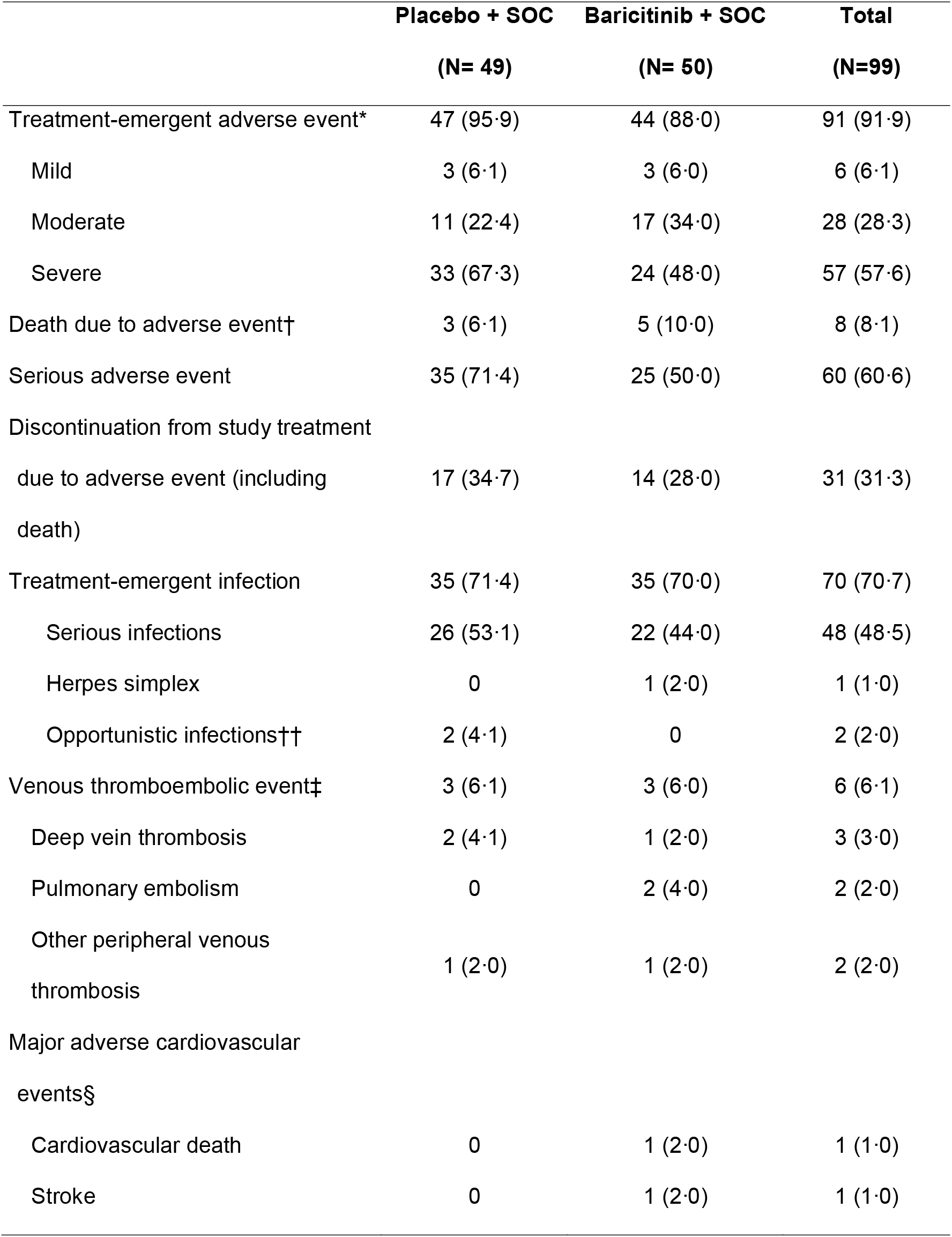
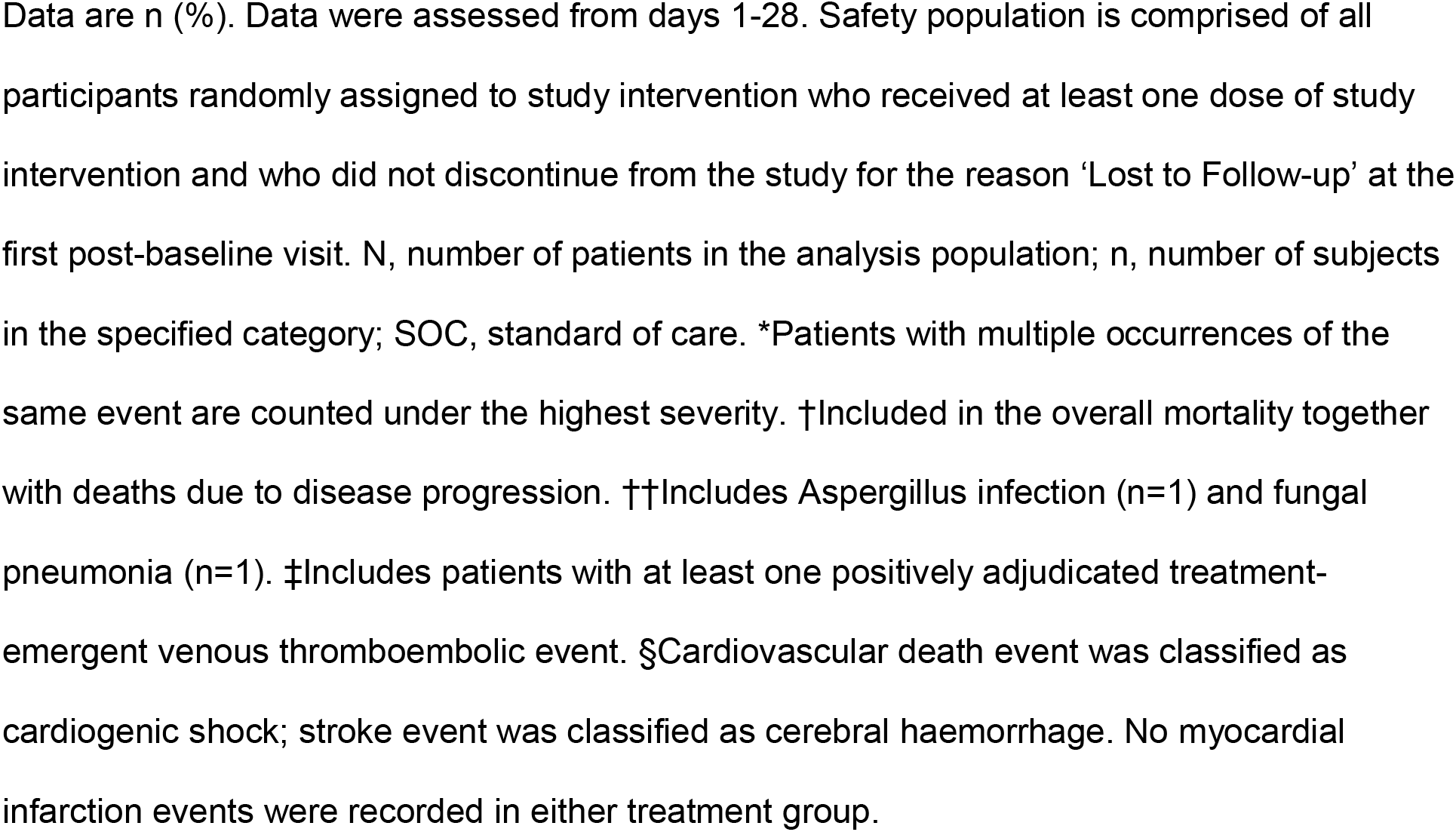
Safety overview in the safety population up to day 28.

## DISCUSSION

This COV-BARRIER addendum trial was a multinational, randomised, double-blind, placebo-controlled trial designed to evaluate the efficacy and safety of baricitinib in participants with COVID-19 who were already receiving IMV or ECMO at enrolment and, to our knowledge, the first study of its kind to evaluate treatment specifically in this patient population with corticosteroids as part of the SOC. Treatment with baricitinib + SOC (including corticosteroids) resulted in an absolute risk reduction of 16·9% in mortality at 60 days (HR 0·56 [95%CI 0·33– 0·97]; p=0·027), which corresponds to a 44% relative reduction in mortality for critically ill participants with COVID-19; overall, one additional death was prevented for every six baricitinib-treated participants at day 28 and day 60. These results are consistent with the reduction in mortality observed in the less severely ill hospitalised participants in the primary COV-BARRIER study population (NIAID-OS 4,5,6) with HR of 0·57 (95% CI 0·41–0·78; p=0·018) resulting in a 43% relative reduction in mortality at day 28 and an absolute risk reduction of 5·0%. As compared to placebo, there was no evidence of an increased risk for infections, serious infections, VTEs, or adverse cardiovascular events in participants treated with baricitinib.

Patients with severe COVID-19 can develop dysregulation of inflammatory mediators such as cytokines interleukin (IL)-6, IL-10, TNF-α, and IFN-γ and chemokines CXCL10 and monocyte chemoattract protein (MCP)-3.^22-24^ Baricitinib, due to the anti-inflammatory action caused by the selective inhibition of JAK1 and JAK2, has been shown to down-regulate these inflammatory mediators implicated in COVID-19 pathophysiology.^25-27^ A macaque study of SARS-CoV-2 infection also found that baricitinib treatment decreased cytokine and chemokine production, reduced the infiltration of inflammatory cells to the lungs, and reduced lung pathology in baricitinib-treated animals, without reducing innate antiviral type 1 INF responses.^28^

The combination of baricitinib with corticosteroids, in particular dexamethasone, may have additive or indeed synergistic effects on these inflammatory molecules for the treatment of patients with COVID-19, as seen by the greater improvement of outcomes when baricitinib and other JAK inhibitors have been studied in combination with SOC which included corticosteroids.^29^ In the current guidelines for the therapeutic management of hospitalised adults with COVID-19 published by the National Institutes of Health (update as of August 25, 2021), baricitinib is recommended in combination with dexamethasone for the treatment of patients who are recently hospitalised and have systemic inflammation and rapidly increasing oxygen requirements.^30^ Despite the recent advances in knowledge of potential treatments for COVID-19, few interventions are included in these guidelines, and options remain limited for patients who require IMV or ECMO. In a recently published meta-analysis of four trials, JAK inhibitors (baricitinib, ruxolitinib, tofacitinib, and nezulcitinib) were reported to reduce mortality risk in patients with COVID-19 by 43% and reduction in risk of requiring IMV or ECMO by 36%;^31^ this analysis does not include the data from COV-BARRIER, which is the first randomised placebo-controlled trial of an immunomodulatory agent to report a reduction in COVID-19 mortality.^32^ The different mortality effect seen in critically ill patients (NIAID-OS7) from COV-BARRIER reported here and ACTT-2^7^ may be driven by the limited use of corticosteroids in ACTT-2 whereas in COV-BARRIER the majority of participants utilised baricitinib in combination with corticosteroids.

Although mortality was significantly reduced following treatment with baricitinib, the overall mortality for those on IMV or ECMO at baseline was 39% and 58% in participants who received baricitinib and placebo, respectively. Published literature provides consistent rates of mortality in this patient population. In one large meta-analysis, the case fatality rate of 45% was reported for this subgroup of patients with COVID-19 requiring IMV with higher mortality rates in older patients and in early epicentres of COVID-19.^5^ RECOVERY, the open-label study which examined the efficacy of the anti-IL-6 antibody tocilizumab for the treatment of patients with COVID-19, reported 28-day mortality of 49% in the participant population receiving tocilizumab while on IMV, vs 51% in the group receiving SOC.^9^

Despite the large effect size seen here in this well conducted, global, randomised, placebo-controlled trial, limitations exist. Limitations include the small sample size (which precludes any firm conclusions regarding other clinical outcomes such as resource utilisation or length of stay), and incomplete control of the SOC, specifically glucocorticoids. In addition, the intensity of these participants hyperinflammatory state may warrant longer durations of immunomodulation that were not part of our study design.

In summary, treatment with baricitinib plus SOC (including use of corticosteroids) in critically ill COVID-19 patients already receiving IMV or ECMO at enrolment resulted in a HR of 0·56, which corresponds to a 44% relative reduction in mortality at 60 days. These results are consistent with the reduction in mortality observed in the less severely ill hospitalised patients in the primary COV-BARRIER study population and further support the use of baricitinib in hospitalised adults with COVID-19. Baricitinib used to treat critically ill persons with COVID-19 represents a novel option to reduce mortality even if the disease process has progressed to the point of already receiving steroids, mechanical ventilation, and ECMO.

## Supporting information

IRB Approval

Redacted Protocol

## Data Availability

Lilly provides access to all individual participant data collected during the trial, after anonymization, with the exception of pharmacokinetic or genetic data. Data are available to request 6 months after the indication studied has been approved in the US and EU and after primary publication acceptance, whichever is later. No expiration date of data requests is currently set once data are made available. Access is provided after a proposal has been approved by an independent review committee identified for this purpose and after receipt of a signed data sharing agreement. Data and documents, including the study protocol, statistical analysis plan, clinical study report, blank or annotated case report forms, will be provided in a secure data sharing environment. For details on submitting a request, see the instructions provided at www.vivli.org.

## Contributors

All authors contributed to the concept and design of the trial, data analysis and interpretation, critical revision of the publication, and final approval to submit, and were accountable for the accuracy and integrity of the publication. CEK, RL, and SC have accessed and verified the underlying data in this report.

## Data sharing

Eli Lilly and Company provides access to all individual participant data collected during the trial, after anonymisation, with the exception of pharmacokinetic or genetic data. Data are available to request 6 months after the indication studied has been approved in the USA and EU or after the trial is completed, whichever is later. No expiration date of data requests is currently set once data are made available. Access is provided after a proposal has been approved by an independent review committee identified for this purpose and after receipt of a signed data sharing agreement. Data and documents (including the study protocol, statistical analysis plan, clinical study report, and blank or annotated case report forms) will be provided in a secure data sharing environment. For details on submitting a request, see the instructions provided at www.vivli.org.

## Acknowledgments

This study was sponsored by Eli Lilly and Company, under license from Incyte Corporation. The authors would like to thank the participants and study investigators and staff, including the COV-BARRIER Study Group who participated in the study, and the following colleagues from Eli Lilly and Company: Anabela Cardoso, David H Adams, and Brenda Crowe for scientific input; and Catherine Lynch for medical writing and process support.

## References

1. Huang C, Wang Y, Li X, et al. Clinical features of patients infected with 2019 novel coronavirus in Wuhan, China. Lancet 2020; 395(10223): 497–506.

2. Ruan Q, Yang K, Wang W, Jiang L, Song J. Clinical predictors of mortality due to COVID-19 based on an analysis of data of 150 patients from Wuhan, China. Intensive Care Med 2020; 46: 846–48.

3. Siddiqi HK, Mehra MR. COVID-19 illness in native and immunosuppressed states: A clinical-therapeutic staging proposal. J Heart Lung Transplant 2020; 39(5): 405–07.

4. Zhou F, Yu T, Du R, et al. Clinical course and risk factors for mortality of adult inpatients with COVID-19 in Wuhan, China: a retrospective cohort study. Lancet 2020; 395: 1054–62.

5. Lim ZJ, Subramaniam A, Ponnapa Reddy M, et al. Case fatality rates for patients with COVID-19 requiring invasive mechanical ventilation. A meta-analysis. Am J Resp Crit Care Med 2021; 203(1): 54–66.

6. Beigel JH, Tomashek KM, Dodd LE, et al. Remdesivir for the treatment of COVID-19 – final report. N Engl J Med 2020; 383(19): 1813–26.

7. Kalil AC, Patterson TF, Mehta AK, et al. Baricitinib plus remdesivir for hospitalized adults with COVID-19. N Engl J Med 2021; 384(9): 795–807.

8. Horby P, Lim WS, Emberson JR, et al. Dexamethasone in hospitalized patients with COVID-19. N Engl J Med 2021; 384(8): 693–704.

9. RECOVERY Collaborative Group. Tocilizumab in patients admitted to hospital with COVID-19 (RECOVERY): a randomised, controlled, open-label, platform trial. Lancet 2021; 397(10285): 1637–45.

10. Goletti D, Cantini F. Baricitinib therapy in COVID-19 pneumonia -an unmet need fulfilled. N Engl J Med 2021; 384(9): 867–69.

11. Marconi VC, Ramanan AV, de Bono S, et al. Efficacy and safety of baricitinib for the treatment of hospitalised adults with COVID-19 (COV-BARRIER): a randomised, double-blind, parallel-group, placebo-controlled phase 3 trial. Lancet Respir Med 2021.

12. Shi JG, Chen X, Lee F, et al. The pharmacokinetics, pharmacodynamics, and safety of baricitinib, an oral JAK 1/2 inhibitor, in healthy volunteers. J Clin Pharmacol 2014; 54(12): 1354–61.

13. McInnes IB, Byers NL, Higgs RE, et al. Comparison of baricitinib, upadacitinib, and tofacitinib mediated regulation of cytokine signaling in human leukocyte subpopulations. Arthritis Res Ther 2019; 21(1): 183.

14. Dörner T, Tanaka Y, Petri MA, et al. Baricitinib-associated changes in global gene expression during a 24-week phase II clinical systemic lupus erythematosus trial implicates a mechanism of action through multiple immune-related pathways. Lupus Sci Med 2020; 7(1): e000424.

15. Richardson P, Griffin I, Tucker C, et al. Baricitinib as potential treatment for 2019-nCoV acute respiratory disease. Lancet 2020; 395(10223): e30–1.

16. Stebbing J, Phelan A, Griffin I, et al. COVID-19: combining antiviral and anti-inflammatory treatments. Lancet Infect Dis 2020; 20: 400–02.

17. Titanji BK, Farley MM, Mehta A, et al. Use of baricitinib in patients with moderate to severe coronavirus disease 2019. Clin Infect Dis 2021; 72(7): 1247–50.

18. Cantini F, Niccoli L, Matarrese D, Nicastri E, Stobbione P, Goletti D. Baricitinib therapy in COVID-19: A pilot study on safety and clinical impact. J Infect 2020; 81(2): 318–56.

19. Cantini F, Niccoli L, Nannini C, et al. Beneficial impact of baricitinib in COVID-19 moderate pneumonia; multicentre study. J Infect 2020; 81(4): 647–79.

20. Stebbing J, Sánchez Nievas G, Falcone M, et al. JAK inhibition reduces SARS-CoV-2 liver infectivity and modulates inflammatory responses to reduce morbidity and mortality. Sci Adv 2021; 7(1): eabe4724.

21. US Food & Drug Administration. Letter of Authorization: EUA for baricitinib (Olumiant) for treatment of coronavirus disease 2019 (COVID-19). 2020. https://www.fda.gov/media/143822/download (accessed August 28 2021).

22. Karki R, Sharma BR, Tuladhar S, et al. Synergism of TNF-α and IFN-γ triggers inflammatory cell death, tissue damage, and mortality in SARS-CoV-2 infection and cytokine shock syndromes. Cell 2021; 184(1): 149–68.e17.

23. Su Y, Chen D, Yuan D, et al. Multi-omics resolves a sharp disease-state shift between mild and moderate COVID-19. Cell 2020; 183(6): 1479–95.e20.

24. Yang Y, Shen C, Li J, et al. Plasma IP-10 and MCP-3 levels are highly associated with disease severity and predict the progression of COVID-19. J Allergy Clin Immunol 2020; 146(1): 119–27.e4.

25. Bronte V, Ugel S, Tinazzi E, et al. Baricitinib restrains the immune dysregulation in patients with severe COVID-19. J Clin Invest 2020; 130(12): 6409–16.

26. Petrone L, Petruccioli E, Alonzi T, et al. In-vitro evaluation of the immunomodulatory effects of baricitinib: implication for COVID-19 therapy. J Infect 2021; 82(4): 58–66.

27. Sims JT, Krishnan V, Chang CY, et al. Characterization of the cytokine storm reflects hyperinflammatory endothelial dysfunction in COVID-19. J Allergy Clin Immunol 2021; 147(1): 107–11.

28. Hoang TN, Pino M, Boddapati AK, et al. Baricitinib treatment resolves lower-airway macrophage inflammation and neutrophil recruitment in SARS-CoV-2-infected rhesus macaques. Cell 2021; 184(2): 460–75.

29. Stebbing J, Lauschke VM. JAK inhibitors – more than just glucocorticoids. N Engl J Med 2021; 385(5): 463–64.

30. COVID-19 treatment guidelines panel. Coronavirus disease 2019 (COVID-19) treatment guidelines. National Institutes of Health. 2021. https://www.covid19treatmentguidelines.nih.gov/ (accessed September 09 2021).

31. Patoulias D, Doumas M, Papadopoulos C, Karagiannis A. Janus kinase inhibitors and major COVID-19 outcomes: time to forget the two faces of Janus! A meta-analysis of randomized controlled trials. Clin Rheumatol 2021.

32. Kalil AC, Stebbing J. Baricitinib: the first immunomodulatory treatment to reduce COVID-19 mortality in a placebo-controlled trial. Lancet Respir Med 2021.

