## Supplementary material for "Baricitinib plus Standard of Care for Hospitalised Adults with COVID-19 on Invasive Mechanical Ventilation or Extracorporeal Membrane Oxygenation: Results of a Randomised, Placebo-Controlled Trial": IRB Approval

B COV-BARRIER OS7 IRBs and Approval Dates

| **Country** | **Site** | **IRB Name** | **Approval Date** |
| --- | --- | --- | --- |
| United States | 102 | South Shore Hospital IRB | 07-Jan-2021 |
| United States | 112 | Western Institutional Review Board - Connexus | 14-Dec-2020 |
| United States | 113 | Western Institutional Review Board - Connexus | 11-Dec-2020 |
| United States | 114 | St. Joseph Health IRB | 09-Dec-2020 |
| United States | 116 | Providence St. Joseph Health | 09-Dec-2020 |
| United States | 133 | Western Institutional Review Board - Connexus | 17-Dec-2020 |
| Mexico | 353 | Instituto Nacional de Cancerologia | 17-Dec-2020 |
| Mexico | 356 | Medica Sur | 07-Dec-2020 |
| Mexico | 357 | Instituto Nacional de Cancerologia | 06-Jan-2021 |
| Argentina | 376 | Stamboulian- Comite de Etica en Investigacion Clínica | 25-Dec-2020 |
| Argentina | 381 | Stamboulian- Comite de Etica en Investigacion Clínica | 25-Dec-2020 |
| Argentina | 383 | Stamboulian- Comite de Etica en Investigacion Clínica | 25-Dec-2020 |
| Brazil | 402 | Hospital Felício Rocho | 26-Mar-2021 |
| Brazil | 409 | Hospital PUC-CAMPINAS | 12-Feb-2021 |
| Brazil | 411 | COMITÊ DE ÉTICA EM PESQUISA DA LIGA NORTE RIOGRANDENSE CONTRA O CANCER | 04-Mar-2021 |
| Brazil | 413 | CEMEC – Centro Multidisciplinar de Estudos Clinicos EPP Ltda | 18-Feb-2021 |
| Brazil | 415 | IPECC - Instituto de Pesquisa Clinica de Campinas | 23-Feb-2021 |
| Brazil | 418 | CECIP - Centro de Estudos do Interior Paulista | 15-Feb-2021 |
