## Supplementary material for "Baricitinib plus Standard of Care for Hospitalised Adults with COVID-19 on Invasive Mechanical Ventilation or Extracorporeal Membrane Oxygenation: Results of a Randomised, Placebo-Controlled Trial": Redacted Protocol

### Title Page

**Confidential Information**

The information contained in this document is confidential and is intended for the use of clinical investigators. It is the property of Eli Lilly and Company or its subsidiaries and should not be copied by or distributed to persons not involved in the clinical investigation of baricitinib (LY3009104), unless such persons are bound by a confidentiality agreement with Eli Lilly and Company or its subsidiaries.

**Note to Regulatory Authorities:** This document may contain protected personal data and/or commercially confidential information exempt from public disclosure. Eli Lilly and Company requests consultation regarding release/redaction prior to any public release. In the United States, this document is subject to Freedom of Information Act (FOIA) Exemption 4 and may not be reproduced or otherwise disseminated without the written approval of Eli Lilly and Company or its subsidiaries.

**Protocol Title:** A Randomized, Double-Blind, Placebo-Controlled, Parallel-Group Phase 3 Study of Baricitinib in Patients with COVID-19 Infection

**Protocol Number:** I4V-MC-KHAA

**Addendum Number:** 5.0

**Addendum Statement:** This addendum is to be performed in addition to all procedures required by protocol I4V-MC-KHAA or any subsequent amendments to that protocol.

**Compound:** baricitinib (LY3009104)

**Sponsor Name:** Eli Lilly and Company
**Legal Registered Address:** Indianapolis, Indiana USA 46285

**Approval Date:** Protocol Addendum (5) Electronically Signed and Approved by Lilly on date provided below.

Approval Date: 01-Dec-2020 GMT

### 1. Rationale for Addendum

This addendum applies to select countries and sites that have the ability to manage patients on invasive mechanical ventilation.

The purpose of this addendum to Protocol I4V-MC-KHAA (KHAA) is to add a cohort of patients that require invasive mechanical ventilation at screening and randomization. Data from this subgroup of patients will inform the risk/benefit profile of baricitinib in patients who are receiving invasive mechanical ventilation or extracorporeal membrane oxygenation (ECMO).

### 2. Protocol Additions

The revised text in this section shows the protocol adjustments implemented by means of this protocol addendum.

Additions to the protocol are designated using underline. Deletions are designated using ~~strikethrough~~.

#### 2.1. Section 3 Objectives and Endpoints

The addition to the protocol is underlined.

| Exploratory |
| --- |
| Exploratory objectives and endpoints may include the following:<br>• C-reactive protein (CRP), D-dimer, lactate dehydrogenase (LDH), ferritin<br>• Virologic measures<br>• Characterization of the pharmacokinetics of baricitinib in intubated patients with COVID-19 infection<br>• Long-term (at least Day 60) clinical outcomes<br>• Evaluate the number of ventilator-free days and mortality through Day 28. |

Note: The Day 28 Clinical Status Assessment is entered for midnight to midnight for the previous day. Therefore, the Day 28 Clinical Status Assessment is entered on Day 29.

#### 2.2. Section 4.2 Scientific Rationale for Study Design

Data from this subgroup of approximately 100 patients will inform the risk/benefit profile of baricitinib in patients who are receiving invasive mechanical ventilation or ECMO at screening and randomization. The addition of the subpopulation of patients requiring invasive mechanical ventilation at baseline is informed from the National Institute of Allergy and Infectious Diseases (NIAID) Adaptive COVID-19 Treatment Trial 2 (ACTT-2).

One hundred and eleven patients receiving invasive mechanical ventilation and/or ECMO at baseline were enrolled in ACTT 2. On Day 15, a larger proportion of patients who received baricitinib + remdesivir showed an improvement in ordinal scale compared to those who received placebo + remdesivir (Figure 1). No new safety signals were detected for baricitinib in the ACTT-2 data.

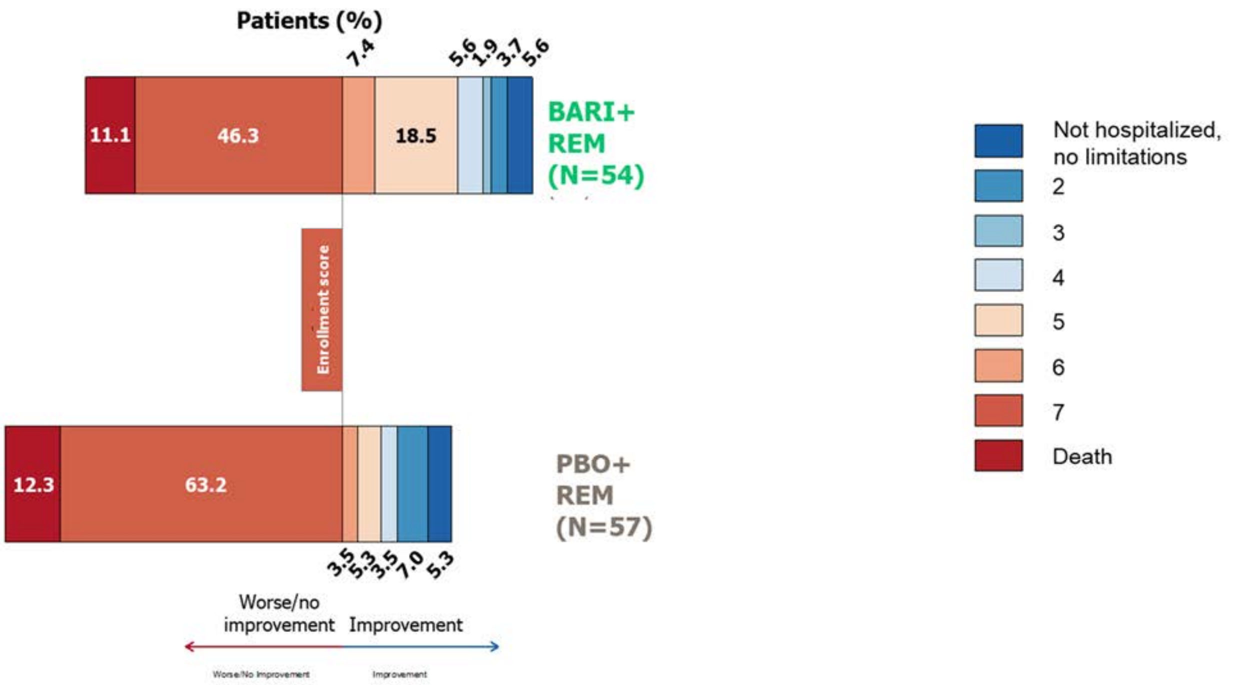

Abbreviations: BARI = baricitinib; PBO = placebo; REM = remdesivir.

**Figure 1. Clinical status scores at Day 15 using an 8-point ordinal scale for patients in Study ACTT-2 who were receiving mechanical ventilation or ECMO at baseline.**

#### 2.3. Section 5 Inclusion and Exclusion Criteria

##### Section 5.1. Inclusion Criteria

[4] Require ~~supplemental oxygen at the time of study entry and at randomization~~ invasive mechanical ventilation or ECMO at screening and randomization.

##### Section 5.2. Exclusion Criteria

~~[13] Require invasive mechanical ventilation, including ECMO at study entry.~~

#### 2.4. Section 6.3 Measures to Minimize Bias: Randomization and Blinding

Patients who meet all criteria for enrollment will be randomized in a 1:1 ratio (baricitinib 4-mg: placebo) at Day 1.

Randomization will be stratified by these factors:

- ~~disease severity:~~
  - ~~hospitalized not requiring supplemental oxygen, requiring ongoing medical care~~
  - ~~hospitalized requiring supplemental oxygen by prongs or mask~~
  - ~~hospitalized requiring non-invasive ventilation or high-flow oxygen~~
- ~~age (<65 years; ≥65 years)~~
- region (United States, Europe, rest of world)
- ~~dexamethasone and/or other systemic corticosteroid used at baseline for primary study condition (Yes/No).~~

#### 2.5. Section 9 Statistical Considerations

##### Section 9.2. Sample Size Determination

The sample size for this cohort is approximately 50 patients per treatment arm and is similar to the number of subjects included in the ACTT-2 study requiring invasive mechanical ventilation or ECMO at baseline. This will approximately double the total number of patients treated with baricitinib in a randomized controlled study, who are hospitalized and on invasive mechanical ventilation or ECMO at baseline.

##### Section 9.4.1. General Considerations

All analyses for this addendum are considered exploratory. These analyses will be conducted separately from the main study cohort of patients in Study KHAA protocol.

Details on the analysis will be defined in the Statistical Analysis Plan (SAP).

**Leo Document ID = 595cd6ac-cec7-4641-ba75-449a09b752c5**

**Approver:** PPD

**Approval Date & Time: 01-Dec-2020 18:54:09 GMT**

**Signature meaning: Approved**

**Approver:** PPD

**Approval Date & Time: 01-Dec-2020 19:15:39 GMT**

**Signature meaning: Approved**
