## Supplemental Appendix for "Baricitinib plus Standard of Care for Hospitalised Adults with COVID-19 on Invasive Mechanical Ventilation or Extracorporeal Membrane Oxygenation: Results of a Randomised, Placebo-Controlled Trial"

### SUPPLEMENTARY APPENDIX

Table of contents

COV-BARRIER Study Group	2

Supplementary Results	3

Mortality	3

Figure S1. Study Design.	4

Table S1: Day 28 all-cause mortality by subgroup	5

Table S2: Ventilator free days through Day 28 by subgroup	7

Table S3: NIAID OS at day 28 by subgroup	8

Table S4: Duration of hospitalisation (days) through Day 28 by subgroup	10

Table S5: Recovery by day 28 by subgroup	11

Table S6: Day 60 all-cause mortality by subgroup	13

Table S7: NIAID OS at day 60 by subgroup	15

Table S8. Follow-up emergent adverse events post day 28 in the safety population	17

#### COV-BARRIER Study Group

**Argentina**

Javier David Altclas, Sanatorio de la Trinidad Mitre, Ciudad Autonoma de Buenos Aires, Buenos Aires

Marcelo Casas, Clinica Adventista Belgrano, Ciudad Autónoma de Buenos Aire, Buenos Aires

Valeria Cevoli Recio, Clinica Viedma, Viedma, Río Negro

**Brazil**

Kleber Giovanni Luz, CPCLIN, Natal, Rio Grande do Norte

Maria Patelli Juliani Souza Lima, Hospital PUC-Campinas, Campinas, SP

Priscila Paulin, CECIP - Centro de Estudos do Interior Paulista, Jaú, SP

Ana Carolina Procopio Carvalho, Hospital Felício Rocho, Belo Horizonte, Minas Gerais

Jose Francisco Kerr Saraiva, IPECC - Instituto de Pesquisa Clinica de Campina, Campinas, SP

Adilson Westheimer Cavalcante, CEMEC – Centro Multidisciplinar de Estudos Clinicos EPP Ltda, São Bernardo do Campo, SP

**Mexico**

Jorge Arturo Alatorre-Alexander, Instituto Nacional de Enfermedades Respiratorias, Mexico City, México

Gustavo Rojas Velasco, Instituto Nacional de Cardiologia Ignacio Chavez, Mexico, DF

Jesus Simon Campos, Hospital General Agustín O'Horán, Yucatan, Merida

**United States**

Todd Ellerin, South Shore Hospital, Weymouth, MA

Jason Goldman, Swedish Medical Center, Seattle, WA

Akram Khan, Oregon Health and Science University, Portland, OR

Imad Shawa, Franciscan St. Francis Health, Indianapolis, IN

Brian Tiffany, Dignity Health Mercy Gilbert Medical Center, Gilbert, AZ

#### Supplementary Results

#### Mortality

At day 28, mortality was reported for 20 of 51 (39·2%) participants in the baricitinib plus SOC group and 29 of 50 (58·0%) participants in the placebo plus SOC group. The HR was 0·54 (95% CI 0·31–0·96) with nominal p=0·030, corresponding to a 46% reduction in the hazard of death. There was an absolute risk reduction of 18·8% and a relative risk reduction of 32·4% in mortality at day 28.

The 60-day mortality remained significantly lower in the baricitinib plus SOC group compared to the placebo plus SOC group. Five additional deaths occurred in the overall population between days 28 and 60, with a similar number of events occuring in both the baricitinib plus SOC (n=3) and placebo plus SOC (n=2) groups. Mortality was reported for 23 of 51 (45·1%) participants in the baricitinib plus SOC group and 31 of 50 (62·0%) participants in the placebo plus SOC group. The HR was 0·56 (95% CI 0·33–0·97) with nominal p=0·027, corresponding to a 44% reduction in the hazard of death. There was an absolute risk reduction of 16·9% and a relative risk reduction of 27·3% in mortality at day 60.

Overall, one additional death was prevented per six baricitinib-treated participants at day 28 and day 60.

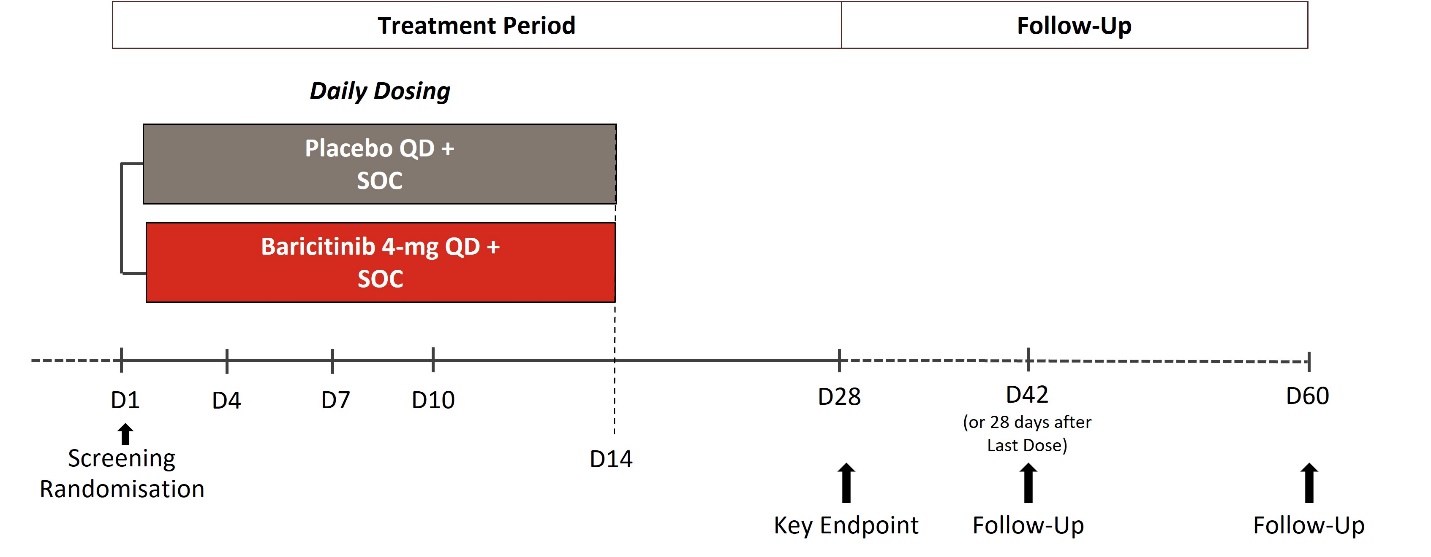

Figure S1. Study Design. Dosing occurred from the day of randomization until day 14, or until hospital discharge or death, whichever comes first. Placebo or baricitinib 4-mg were given in addition to SOC as per local clinical practice for management of COVID 19, as defined in the protocol. D=study day. QD=once-daily. SOC=standard of care.

#### Table S1: Day 28 all-cause mortality by subgroup

|  | **Placebo + SOC**  **(N=50)** | **Baricitinib + SOC**  **(N=51)** | **Comparison with**  **placebo (95% CI)** | **Nominal p value*** |
| --- | --- | --- | --- | --- |
| Baseline corticosteroid use | N-obs=44 | N-obs=43 |  |  |
| n (%)‡ | 26 (59·1) | 16 (37·2) | 0·48 (0·26, 0·91) | 0·023 |
| KM Estimates (95% CI) | 60·4 (41·1, 80·2) | 38·8 (23·2, 59·8) |  |  |
| Time to mortality, days; median (95% CI) | 17·0 (11·0, NA) | NA (24·0, NA) |  |  |
| No baseline corticosteroid use | N-obs=6 | N-obs=8 |  |  |
| n (%)‡ | 3 (50·0) | 4 (50·0) | 0·22 (0, 4·15) | 0·75 |
| KM Estimates (95% CI) | 50·0 (14·4, 95·5) | 50·0 (17·2, 92·1) |  |  |
| Time to mortality, days; median (95% CI) | NA (6·0, NA) | NA (7·0, NA) |  |  |
| Baseline remdesivir use | N-obs=2 | N-obs=0 |  |  |
| n (%)‡ | 0 (0) | - | NA | - |
| KM Estimates (95% CI) | - | - |  |  |
| Time to mortality, days; median (95% CI) | NA (NA, NA) | - |  |  |
| No baseline remdesivir use | N-obs=48 | N-obs=51 |  |  |
| n (%)‡ | 29 (60·4) | 20 (39·2) | 0·22 (0, 4·15) | 0·016 |
| KM Estimates (95% CI) | 50·0 (14·4, 95·5) | 40·6 (25·8, 59·7) |  |  |
| Time to mortality, days; median (95% CI) | 16·0 (11·0, NA) | NA (24·0, NA) |  |  |
| Country: Argentina | N-obs=9 | N-obs=12 |  |  |
| n (%)‡ | 8 (88·9) | 8 (66·7) | 0·42 (0·14, 1·26) | 0·026 |
| KM Estimates (95% CI) | 88·9 (32·2, 100·0) | 66·7 (31·7, 95·8) |  |  |
| Time to mortality, days; median (95% CI) | 8·0 (3·0, 20·0) | 22·5 (8·0, NA) |  |  |
| Country: Brazil | N-obs=14 | N-obs=15 |  |  |
| n (%)‡ | 5 (35·7) | 5 (33·3) | 0·92 (0·26, 3·22) | 0·99 |
| KM Estimates (95% CI) | 35·7 (13·9, 73·0) | 35·4 (13·6, 72·8) |  |  |
| Time to mortality, days; median (95% CI) | NA (10·0, NA) | NA (9·0, NA) |  |  |
| Country: Mexico | N-obs=17 | N-obs=14 |  |  |
| n (%)‡ | 10 (58·8) | 4 (28·6) | 0·33 (0·10, 1·08) | 0·048 |
| KM Estimates (95% CI) | 63·0 (31·7, 92·6) | 28·6 (9·8, 66·7) |  |  |
| Time to mortality, days; median (95% CI) | 19·0 (8·0, NA) | NA (20·0, NA) |  |  |
| Country: United States | N-obs=10 | N-obs=10 |  |  |
| n (%)‡ | 6 (60·0) | 3 (30·0) | 0·46 (0·11, 1·84) | 0·29 |
| KM Estimates (95% CI) | 60·0 (25·4, 94·3) | 33·3 (9·5, 80·7) |  |  |
| Time to mortality, days; median (95% CI) | 15·0 (4·0, NA) | NA (5·0, NA) |  |  |

Data are presented as mean (SD) unless otherwise specified. Data were assessed from days 1-28. Hazard ratios, 95% CIs, and p values are calculated using Cox proportional hazard regression model adjusted for age (<65 years, ≥65 years) and region (United States, rest of world). p values are for comparisons of baricitinib 4-mg with placebo. *All endpoints are exploratory due to the nature of the study. Nominal p-values are shown. ‡Comparisons are hazard ratio. CI, confidence interval; KM, Kaplan-Meier; N, number of patients in the analysis population; N-obs=number of participants in the analysis; n, number of subjects in the specified category; NA, not applicable; SOC, standard of care.

#### Table S2: Ventilator free days through Day 28 by subgroup

|  | **Placebo + SOC**  **(N=50)** | **Baricitinib + SOC**  **(N=51)** | **Comparison with**  **placebo (95% CI)** | **Nominal p value*** |
| --- | --- | --- | --- | --- |
| Baseline corticosteroid use | N-obs=44 | N-obs=43 |  |  |
|  | 5·2 (8·0) | 8·3 (9·9) | 2·94 (–0·83, 6·72) | 0·12 |
| No baseline corticosteroid use | N-obs=6 | N-obs=8 |  |  |
|  | 7·5 (11·6) | 6·8 (12·5) | 5·05 (–12·03, 22·12) | 0·52 |
| Baseline remdesivir use | N-obs=2 | N-obs=0 |  |  |
|  | 20·5 (2·1) | - | - | - |
| No baseline remdesivir use | N-obs=48 | N-obs=51 |  |  |
|  | 4·9 (8·0) | 8·1 (10·2) | 3·01 (–0·69, 6·71) | 0·11 |
| Country: Argentina | N-obs=9 | N-obs=12 |  |  |
|  | 2·7 (8·0) | 4·9 (8·5) | –1·67 (–9·71, 6·38) | 0·67 |
| Country: Brazil | N-obs=14 | N-obs=15 |  |  |
|  | 6·6 (8·1) | 10·3 (11·6) | 3·34 (–4·42, 11·11) | 0·38 |
| Country: Mexico | N-obs=17 | N-obs=14 |  |  |
|  | 5·5 (8·5) | 8·6 (10·2) | 3·14 (–3·86, 10·15) | 0·37 |
| Country: United States | N-obs=10 | N-obs=10 |  |  |
|  | 6·4 (9·5) | 7·7 (10·6) | 2·75 (–6·49, 11·99) | 0·54 |

Data are presented as mean (SD). Data were assessed from days 1-28. p values and 95% CI are calculated using ANOVA model adjusted for age (<65 years, ≥65 years) and region (United States, rest of world) for comparisons of baricitinib 4-mg with placebo. *All endpoints are exploratory due to the nature of the study. Nominal p-values are shown. CI, confidence interval; N, number of patients in the analysis population; N-obs=number of participants in the analysis; n, number of subjects in the specified category; SOC, standard of care.

#### Table S3: NIAID OS at day 28 by subgroup

|  | **Placebo + SOC**  **(N=50)** | **Baricitinib + SOC**  **(N=51)** | **Total**  **(N=101)** |
| --- | --- | --- | --- |
| Overall | N-obs=50 | N-obs=51 | N-obs=101 |
| OS1 | 7 (14·0) | 10 (19·6) | 17 (16·8) |
| OS2 | 6 (12·0) | 9 (17·6) | 15 (14·9) |
| OS3 | 0 | 0 | 0 |
| OS4 | 0 | 0 | 0 |
| OS5 | 0 | 4 (7·8) | 4 (4·0) |
| OS6 | 0 | 0 | 0 |
| OS7 | 6 (12·0) | 6 (11·8) | 12 (11·9) |
| OS8 | 29 (58·0) | 20 (39·2) | 49 (48·5) |
| Missing | 2 (4·0) | 2 (3·9) | 4 (4·0) |
| Baseline corticosteroid use | N-obs=44 | N-obs=43 | N-obs=87 |
| OS1 | 6 (13·6) | 8 (18·6) | 14 (16·1) |
| OS2 | 5 (11·4) | 9 (20·9) | 14 (16·1) |
| OS3 | 0 | 0 | 0 |
| OS4 | 0 | 0 | 0 |
| OS5 | 0 | 4 (9·3) | 4 (4·6) |
| OS6 | 0 | 0 | 0 |
| OS7 | 5 (11·4) | 4 (9·3) | 4 (4·6) |
| OS8 | 26 (59·1) | 16 (37·2) | 42 (48·3) |
| Missing | 2 (4·5) | 2 (4·7) | 4 (4·6) |
| No baseline corticosteroid use | N-obs=6 | N-obs=8 | N-obs=14 |
| OS1 | 1 (16·7) | 2 (25·0) | 3 (21·4) |
| OS2 | 1 (16·7) | 0 | 1 (7·1) |
| OS3 | 0 | 0 | 0 |
| OS4 | 0 | 0 | 0 |
| OS5 | 0 | 0 | 0 |
| OS6 | 0 | 0 | 0 |
| OS7 | 1 (16·7) | 2 (25·0) | 3 (21·4) |
| OS8 | 3 (50·0) | 4 (50·0) | 7 (50·0) |
| Missing | 0 | 0 | 0 |
| Baseline remdesivir use | N-obs=2 | N-obs=0 | N-obs=2 |
| OS1 | 0 | 0 | 0 |
| OS2 | 2 (100·0) | 0 | 2 (100·0) |
| OS3 | 0 | 0 | 0 |
| OS4 | 0 | 0 | 0 |
| OS5 | 0 | 0 | 0 |
| OS6 | 0 | 0 | 0 |
| OS7 | 0 | 0 | 0 |
| OS8 | 0 | 0 | 0 |
| Missing | 0 | 0 | 0 |
| No baseline remdesivir use | N-obs=48 | N-obs=51 | N-obs=99 |
| OS1 | 7 (14·6) | 10 (19·6) | 17 (17·2) |
| OS2 | 4 (8·3) | 9 (17·6) | 13 (13·1) |
| OS3 | 0 | 0 | 0 |
| OS4 | 0 | 0 | 0 |
| OS5 | 0 | 4 (7·8) | 4 (4·0) |
| OS6 | 0 | 0 | 0 |
| OS7 | 6 (12·5) | 6 (11·8) | 12 (12·1) |
| OS8 | 29 (60·4) | 20 (39·2) | 49 (49·5) |
| Missing | 2 (4·2) | 2 (3·9) | 4 (4·0) |

Data are presented as n (%). Data were assessed from days 1-28. Percentage of response is calculated by responder/n*100. N, number of patients in the analysis population; N-obs=number of participants in the analysis; n, number of subjects in the specified category; SOC, standard of care.

#### Table S4: Duration of hospitalisation (days) through Day 28 by subgroup

|  | **Placebo + SOC**  **(N= 50)** | **Baricitinib + SOC**  **(N= 51)** | **Comparison with**  **placebo (95% CI)** | **Nominal p value*** |
| --- | --- | --- | --- | --- |
| Baseline corticosteroid use | N-obs=44 | N-obs=43 |  |  |
|  | 26·3 (3·5) | 23·9 (6·7) | –2·36 (–4·63, -0·09) | 0·042 |
| No baseline corticosteroid use | n=6 | n=8 |  |  |
|  | 24·5 (6·4) | 23·0 (9·3) | –5·11 (–16·07, 5·84) | 0·32 |
| Baseline remdesivir use | N-obs=2 | N-obs=0 |  |  |
|  | 21·0 (2·8) | - | - | - |
| No baseline remdesivir use | N-obs=48 | N-obs=51 |  |  |
|  | 26·3 (3·8) | 23·7 (7·1) | –2·54 (–4·86, –0·21) | 0·033 |
| Country: Argentina | N-obs=9 | N-obs=12 |  |  |
|  | 26·3 (5·0) | 26·6 (4·9) | 1·67 (–3·52, 6·85) | 0·51 |
| Country: Brazil | N-obs=14 | N-obs=15 |  |  |
|  | 25·6 (3·4) | 22·2 (8·2) | –3·16 (–8·08, 1·77) | 0·20 |
| Country: Mexico | N-obs=17 | N-obs=14 |  |  |
|  | 26·4 (4·1) | 22·8 (7·5) | –3·56 (–7·98, 0·86) | 0·11 |
| Country: United States | N-obs=10 | N-obs=10 |  |  |
|  | 25·9 (3·51) | 23·9 (6·7) | –2·83 (–7·69, 2·02) | 0·24 |

Data are presented as mean (SD). Data were assessed from days 1-28. p values and 95% CI are calculated using ANOVA model adjusted for age (<65 years, ≥65 years) and region (United States, rest of world) for comparisons of baricitinib 4-mg with placebo. *All endpoints are exploratory due to the nature of the study. Nominal p-values are shown. CI, confidence interval; N, number of patients in the analysis population; N-obs=number of participants in the analysis; n, number of subjects in the specified category; SOC, standard of care.

#### Table S5: Recovery by day 28 by subgroup

|  | **Placebo + SOC**  **(N=50)** | **Baricitinib + SOC**  **(N=51)** | **Comparison with**  **placebo (95% CI)** | **Nominal p value*** |
| --- | --- | --- | --- | --- |
| Baseline corticosteroid use | N-obs=44 | N-obs=43 |  |  |
| n (%)‡ | 11 (25·0) | 17 (39·5) | 1·85 (0·86, 3·95) | 0·10 |
| KM Estimates (95% CI) | 26·2 (13·8, 46·1) | 41·4 (18·9, 56·4) |  |  |
| Time to recovery, days; median (95% CI) | NA (NA, NA) | NA (25·0, NA) |  |  |
| No baseline corticosteroid use | N-obs=6 | N-obs=8 |  |  |
| n (%)‡ | 2 (33·3) | 2 (25·0) | 2·37 (0·23, 24·44) | 0·88 |
| KM Estimates (95% CI) | 33·3 (7·2, 89·1) | 25·0 (5·3, 77·9) |  |  |
| Time to recovery, days; median (95% CI) | NA (13·0, NA) | NA (8·0, NA) |  |  |
| Baseline remdesivir use | N-obs=2 | N-obs=0 |  |  |
| n (%)‡ | 2 (100) | - | NA | - |
| KM Estimates (95% CI) | 100·0 (NA, NA) | - |  |  |
| Time to recovery, days; median (95% CI) | 22·0 (20·0, NA) | - |  |  |
| No baseline remdesivir use | N-obs=48 | N-obs=51 |  |  |
| n (%)‡ | 11 (22·9) | 19 (37·3) | 1· 80 (0·86, 3·80) | 0·083 |
| KM Estimates (95% CI) | 23· 8 (12·5, 42·4) | 38· 7 (18·8, 52·6) |  |  |
| Time to recovery, days; median (95% CI) | NA (NA, NA) | NA (28·0, NA) |  |  |
| Country: Argentina | N-obs=9 | N-obs=12 |  |  |
| n (%)‡ | 1 (11·1) | 1 (8·3) | 0· 0 (0, NA) | 0·86 |
| KM Estimates (95% CI) | 11· 1 (NA, NA) | 8· 3 (NA, NA) |  |  |
| Time to recovery, days; median (95% CI) | NA (14·0, NA) | NA (NA, NA) |  |  |
| Country: Brazil | N-obs=14 | N-obs=15 |  |  |
| n (%)‡ | 6 (42·9) | 8 (53·3) | 1· 43 (0·49, 4·16) | 0·35 |
| KM Estimates (95% CI) | 42· 9 (18·3, 78·9) | 56· 67 (16·7, NA) |  |  |
| Time to recovery, days; median (95% CI) | NA (25·0, NA) | 28· 0 (12·0, NA) |  |  |
| Country: Mexico | N-obs=17 | N-obs=14 |  |  |
| n (%)‡ | 3 (17·6) | 6 (42·9) | 2· 74 (0·68, 11·01) | 0·14 |
| KM Estimates (95% CI) | 19· 3 (5·4, 56·3) | 42· 9 (13·1, 75·2) |  |  |
| Time to recovery, days; median (95% CI) | NA (NA, NA) | NA (15·0, NA) |  |  |
| Country: United States | N-obs=10 | N-obs=10 |  |  |
| n (%)‡ | 3 (30·0) | 4 (40·0) | 2· 55 (0·53, 12·28) | 0·51 |
| KM Estimates (95% CI) | 30· 0 (8·6, 75·6) | 44· 4 (15·2, 87·8) |  |  |
| Time to recovery, days; median (95% CI) | NA (20·0, NA) | NA (9·0, NA) |  |  |

Data are presented as mean (SD) unless otherwise specified. Data were assessed from days 1-28. The rate ratio, 95% CI, and p value was calculated using a Cox proportional hazards model adjusted for age (<65 years, ≥65 years) and region (United States, rest of world). p values are for comparisons of baricitinib 4-mg with placebo. *All endpoints are exploratory due to the nature of the study. Nominal p-values are shown. ‡Comparisons are hazard ratio. CI, confidence interval; KM, Kaplan-Meier; N, number of patients in the analysis population; N-obs=number of participants in the analysis; n, number of subjects in the specified category; NA, not applicable; SOC, standard of care.

#### Table S6: Day 60 all-cause mortality by subgroup

|  | **Placebo + SOC**  **(N=50)** | **Baricitinib + SOC**  **(N=51)** | **Comparison with**  **placebo (95% CI)** | **Nominal p value*** |
| --- | --- | --- | --- | --- |
| Overall | N-obs=50 | N-obs=51 |  |  |
| n (%)‡ | 31 (62·0) | 23 (45·1) | 0·56 (0·33, 0·97) | 0·027 |
| KM Estimates (95% CI) | 63·3 (44·7, 81·7) | 45·4 (30·0, 64·2) |  |  |
| Time to mortality, days; median (95% CI) | 17·0 (11·0, NA) | NA (24·0, NA) |  |  |
| Baseline corticosteroid use | N-obs=44 | N-obs=43 |  |  |
| n (%)‡ | 28 (63·6) | 19 (44·2) | 0·50 (0·28, 0·90) | 0·020 |
| KM Estimates (95% CI) | 65·3 (45·1, 84·6) | 44·4 (28·0, 64·9) |  |  |
| Time to mortality, days; median (95% CI) | 17·0 (11·0, 43·0) | NA (25·0, NA) |  |  |
| No baseline corticosteroid use | N-obs=6 | N-obs=8 |  |  |
| n (%)‡ | 3 (50·0) | 4 (50·0) | 0·22 (0, 4·15) | 0·75 |
| KM Estimates (95% CI) | 50·0 (14·4, 95·5) | 50·0 (17·2, 92·1) |  |  |
| Time to mortality, days; median (95% CI) | NA (6·0, NA) | NA (7·0, NA) |  |  |
| Baseline remdesivir use | N-obs=2 | N-obs=0 |  |  |
| n (%)‡ | 0 (0) | - | NA | - |
| KM Estimates (95% CI) | - | - |  |  |
| Time to mortality, days; median (95% CI) | NA (NA, NA) | - |  |  |
| No baseline remdesivir use | N-obs=48 | N-obs=51 |  |  |
| n (%)‡ | 31 (64·6) | 23 (45·1) | 0·52 (0·30, 0·90) | 0·013 |
| KM Estimates (95% CI) | 66·1 (46·6, 84·5) | 45·4 (30·0, 64·2) |  |  |
| Time to mortality, days; median (95% CI) | 16·0 (11·0, 30·0) | NA (24·0, NA) |  |  |
| Country: Argentina | N-obs=9 | N-obs=12 |  |  |
| n (%)‡ | 8 (88·9) | 8 (66·7) | 0·42 (0·14, 1·26) | 0·026 |
| KM Estimates (95% CI) | 88·9 (32·2, 100·0) | 66·7 (31·7, 95·8) |  |  |
| Time to mortality, days; median (95% CI) | 8·0 (3·0, 20·0) | 22·5 (8·0, NA) |  |  |
| Country: Brazil | N-obs=14 | N-obs=15 |  |  |
| n (%)‡ | 7 (50·0) | 5 (33·3) | 0·70 (0·21, 2·27) | 0·46 |
| KM Estimates (95% CI) | 50·0 (22·9, 84·3) | 33·3 (12·9, 69·6) |  |  |
| Time to mortality, days; median (95% CI) | NA (10·0, NA) | NA (9·0, NA) |  |  |
| Country: Mexico | N-obs=17 | N-obs=14 |  |  |
| n (%)‡ | 10 (58·8) | 6 (42·9) | 0·47 (0·17, 1·31) | 0·14 |
| KM Estimates (95% CI) | 63·0 (31·7, 92·6) | 44·9 (18·6, 82·2) |  |  |
| Time to mortality, days; median (95% CI) | 19·0 (8·0, NA) | NA (20·0, NA) |  |  |
| Country: United States | N-obs=10 | N-obs=10 |  |  |
| n (%)‡ | 6 (60·0) | 4 (40·0) | 0·50 (0·14, 1·80) | 0·35 |
| KM Estimates (95% CI) | 60·0 (25·4, 94·3) | 40·0 (13·8, 82·8) |  |  |
| Time to mortality, days; median (95% CI) | 15·0 (4·0, NA) | NA (5·0, NA) |  |  |

Data are presented as mean (SD) unless otherwise specified. Data were assessed from days 1-60. Hazard ratios, 95% CIs, and p values are calculated using Cox proportional hazard regression model adjusted for age (<65 years, ≥65 years) and region (United States, rest of world) for comparisons of baricitinib 4-mg with placebo. *All endpoints are exploratory due to the nature of the study. Nominal p-values are shown. ‡Comparisons are hazard ratio. CI, confidence interval; KM, Kaplan-Meier; N, number of patients in the analysis population; N-obs=number of participants in the analysis; n, number of subjects in the specified category; SOC, standard of care.

#### Table S7: NIAID OS at day 60 by subgroup

|  | **Placebo + SOC**  **(N=50)** | **Baricitinib + SOC**  **(N=51)** | **Total**  **(N=101)** |
| --- | --- | --- | --- |
| Overall | N-obs=50 | N-obs=51 | N-obs=101 |
| OS1 | 11 (22·0) | 16 (31·4) | 27 (26·7) |
| OS2 | 5 (10·0) | 8 (15·7) | 13 (12·9) |
| OS3 | 0 | 0 | 0 |
| OS4 | 0 | 0 | 0 |
| OS5 | 0 | 0 | 0 |
| OS6 | 0 | 0 | 0 |
| OS7 | 1 (2·0) | 1 (2·0) | 2 (2·0) |
| OS8 | 31 (62·0) | 23 (45·1) | 54 (53·5) |
| Missing | 2 (4·0) | 3 (5·9) | 5 (5·0) |
| Baseline corticosteroid use | N-obs=44 | N-obs=43 | N-obs=87 |
| OS1 | 9 (20·5) | 14 (32·6) | 23 (26·4) |
| OS2 | 4 (9·1) | 8 (18·6) | 12 (13·8) |
| OS3 | 0 | 0 | 0 |
| OS4 | 0 | 0 | 0 |
| OS5 | 0 | 0 | 0 |
| OS6 | 0 | 0 | 0 |
| OS7 | 1 (2·3) | 0 | 1 (1·1) |
| OS8 | 28 (63·6) | 19 (44·2) | 47 (54·0) |
| Missing | 2 (4·5) | 2 (4·7) | 4 (4·6) |
| No baseline corticosteroid use | N-obs=6 | N-obs=8 | N-obs=14 |
| OS1 | 2 (33·3) | 2 (25·0) | 4 (28·6) |
| OS2 | 1 (16·7) | 0 | 1 (7·1) |
| OS3 | 0 | 0 | 0 |
| OS4 | 0 | 0 | 0 |
| OS5 | 0 | 0 | 0 |
| OS6 | 0 | 0 | 0 |
| OS7 | 0 | 1 (12·5) | 1 (7·1) |
| OS8 | 3 (50·0) | 4 (50·0) | 7 (50·0) |
| Missing | 0 | 1 (12·5) | 1 (7·1) |
| Baseline remdesivir use | N-obs=2 | N-obs=0 | N-obs=2 |
| OS1 | 1 (50·0) | 0 | 1 (50·0) |
| OS2 | 1 (50·0) | 0 | 1 (50·0) |
| OS3 | 0 | 0 | 0 |
| OS4 | 0 | 0 | 0 |
| OS5 | 0 | 0 | 0 |
| OS6 | 0 | 0 | 0 |
| OS7 | 0 | 0 | 0 |
| OS8 | 0 | 0 | 0 |
| Missing | 0 | 0 | 0 |
| No baseline remdesivir use | N-obs=48 | N-obs=51 | N-obs=99 |
| OS1 | 10 (20·8) | 16 (31·4) | 26 (26·3) |
| OS2 | 4 (8·3) | 8 (15·7) | 12 (12·1) |
| OS3 | 0 | 0 | 0 |
| OS4 | 0 | 0 | 0 |
| OS5 | 0 | 0 | 0 |
| OS6 | 0 | 0 | 0 |
| OS7 | 1 (2·1) | 1 (2·0) | 2 (2·0) |
| OS8 | 31 (64·6) | 23 (45·1) | 54 (54·5) |
| Missing | 2 (4·2) | 3 (5·9) | 5 (5·1) |

Data are presented as n (%). Data were assessed from days 1-60. Percentage of response is calculated by responder/n*100. N, number of patients in the analysis population; N-obs=number of participants in the analysis; n, number of subjects in the specified category; SOC, standard of care.

#### Table S8. Follow-up emergent adverse events post day 28 in the safety population

|  | **Placebo + SOC**  **(N= 49)** | **Baricitinib + SOC**  **(N= 50)** | **Total**  **(N=99)** |
| --- | --- | --- | --- |
| Follow-up emergent adverse event* | 4 (8·2) | 5 (10·0) | 9 (9·1) |
| Infections | 0 | 3 (6·0) | 3 (3·0) |
| Venous thromboembolic event‡ | 0 | 1 (3·3) | 1 (2·0) |
| Deep vein thrombosis | 0 | 1 (3·3) | 1 (2·0) |
| Pulmonary embolism | 0 | 0 | 0 |
| Other peripheral venous thrombosis | 0 | 0 | 0 |

Data are n (%). Data were assessed from days 28-60. Safety population is comprised of all participants randomly assigned to study intervention who received at least 1 dose of study intervention and who did not discontinue from the study for the reason ‘Lost to Follow-up’ at the first postbaseline visit. N, number of patients in the analysis population; n, number of subjects in the specified category; SOC, standard of care. *Participants with ≥1 emergent adverse event. ‡Includes patients with at least one positively adjudicated venous thromboembolic event. The number of participants in the follow-up analysis set was as follows: Placebo + SOC N=19; Baricitinib + SOC N=30.
